## Supplementary material 1 for "How effective were Australian Quarantine Stations in mitigating transmission aboard ships during the influenza pandemic of 1918-19?"

### How effective were Australian Quarantine Stations in mitigating transmission aboard ships during the influenza pandemic of 1918-19?
### Supplementary Material 1
### Modelling and assumptions

Punya Alahakoon[1,3,4], Peter G. Taylor[1], James M. McCaw[*,1,2]

[1]School of Mathematics and Statistics , The University of Melbourne, Melbourne, Australia.
[2]Centre for Epidemiology and Biostatistics, Melbourne School of Population and Global Health, The University of Melbourne, Melbourne, Australia.
[3]School of Population Health, University of New South Wales, Sydney, Australia.
[4]Kirby Institute, University of New South Wales, Sydney, Australia.

#### S1 Data

##### S1.1 Availability of the ship outbreak data

The data and the related assumptions we made for the outbreaks on board *Medic, Boonah, Devon,* and *Manuka* are included as separate files and they are available on GitHub. Please refer to the link `https://github.com/PunyaAlahakoon/Ship_outbreaks_1918.git`.

[*]

#### S1.2 Snapshots of ship outbreaks

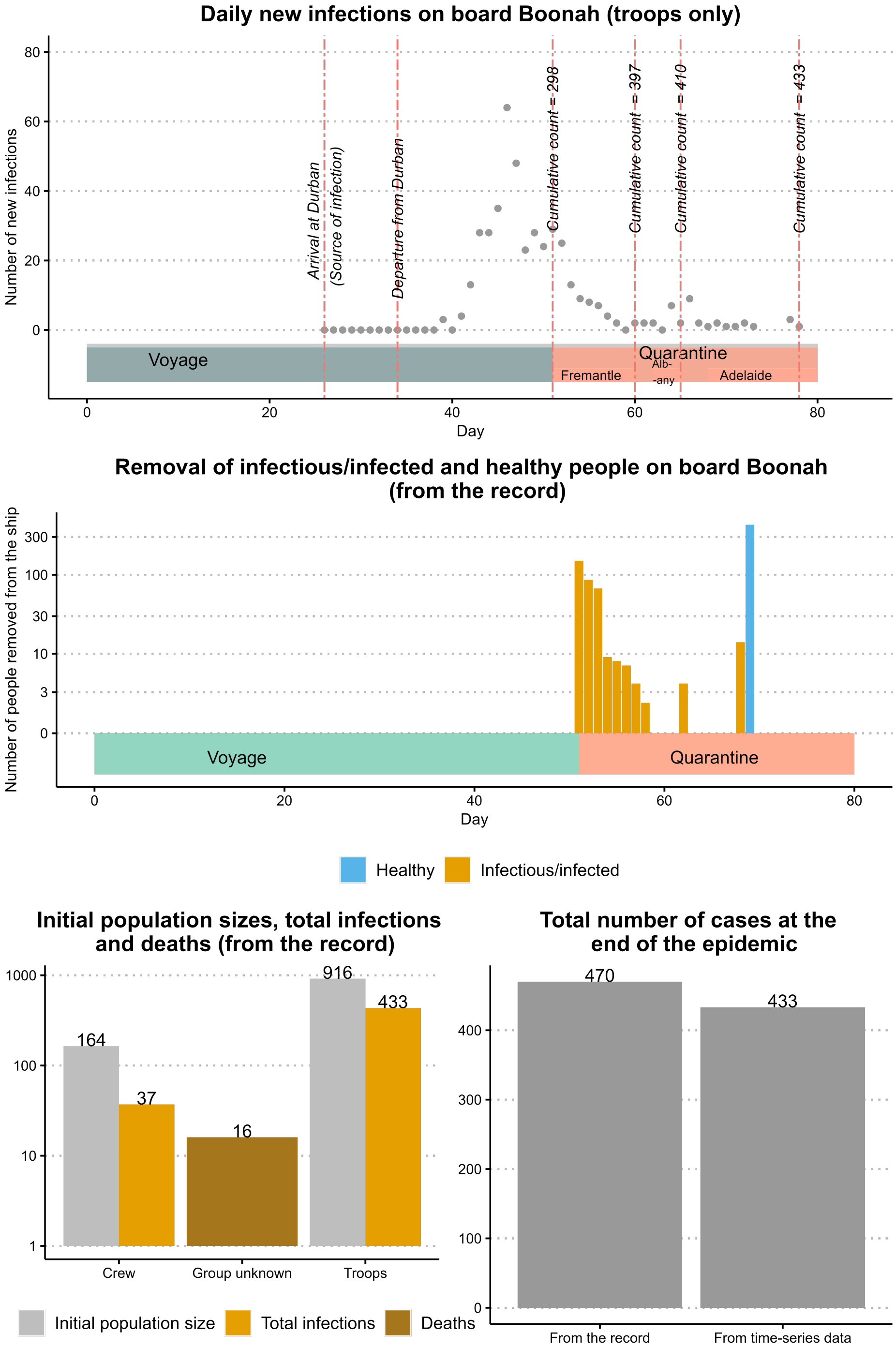


Figure S1: A snapshot of data relating to the outbreak on board *Boonah.*
The top panel display the daily new infections among the troops during the voyage and quarantine period in Fremantle. The middle panel displays the removal of infectious/ infected and healthy people on board *Boonah* during the quarantine period. The bottom compares the total infections and deaths that occurred with respect to the initial population size by group.

Table S1: Removal of healthy, infectious, and infected individuals from the *Boonah*

| **Day** | **Healthy removals** | **Total infectious/infected Removals** |
|---|---|---|
| 51 | 0 | 150 |
| 52 | 0 | 86 |
| 53 | 0 | 67 |
| 54 | 0 | 9 |
| 55 | 0 | 8 |
| 56 | 0 | 7 |
| 57 | 0 | 4 |
| 58 | 0 | 2 |
| 62 | 0 | 4 |
| 68 | 0 | 14 |
| 69 | 427 | 0 |

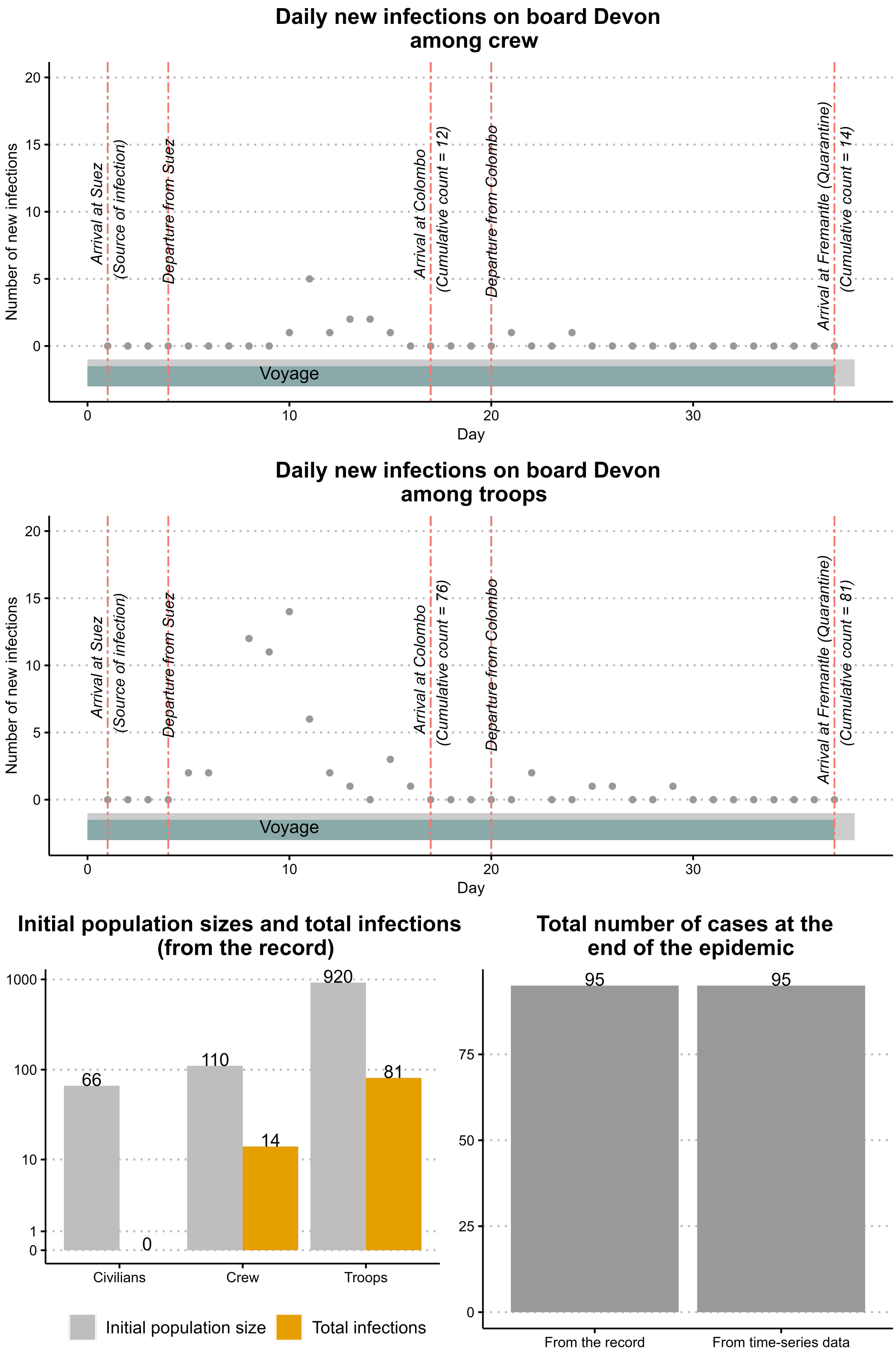


Figure S2: A snapshot of data relating to the outbreak on board *Devon*. The top panel displays the daily new infections among the crew of *Devon* during the voyage. The middle panel displays the daily new infections among the troops of *Devon* during the voyage. The bottom compares the total infections and deaths that occurred with respect to the initial population size by group.

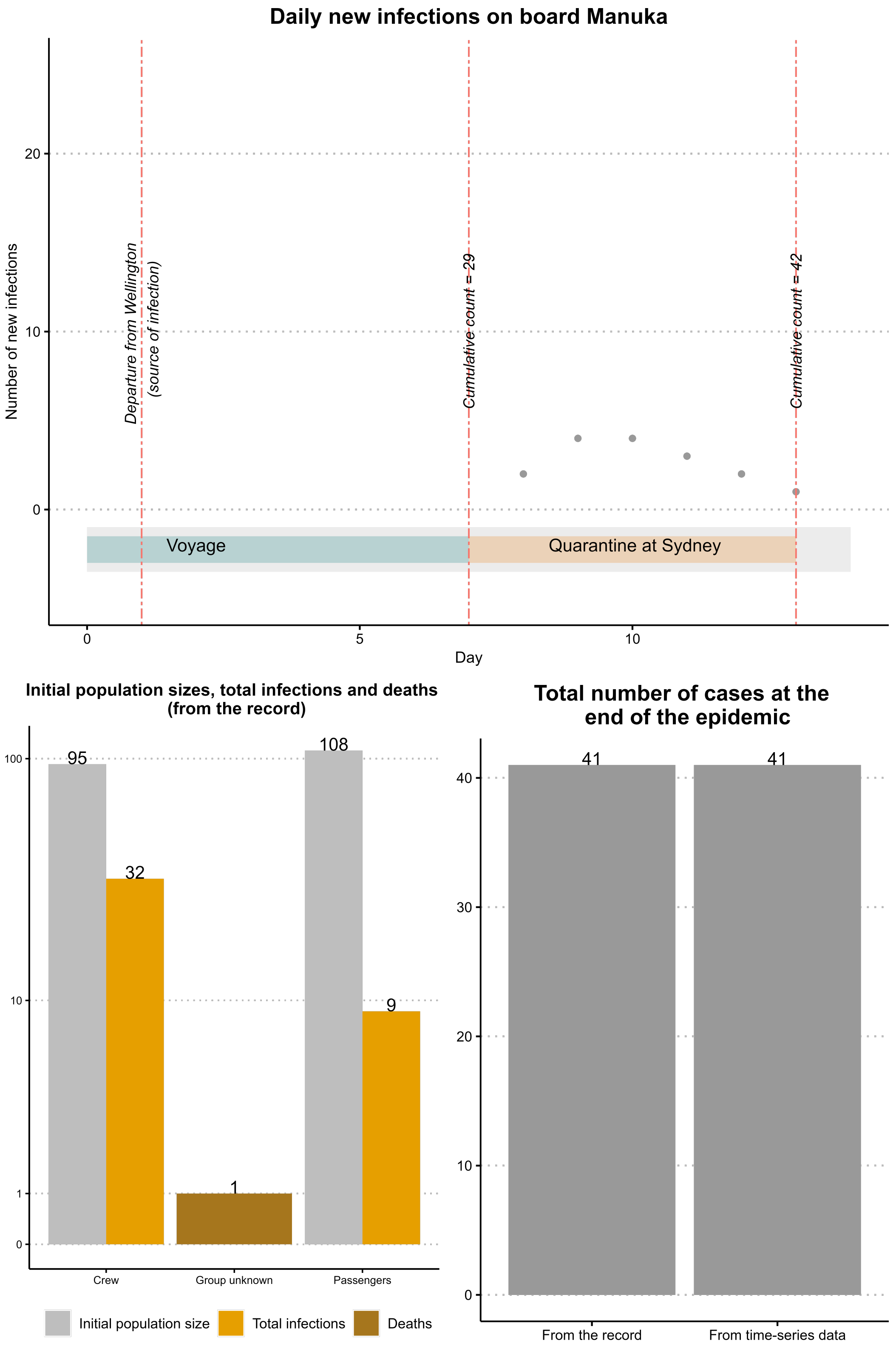


Figure S3: A snapshot of data relating to the outbreak on board *Manuka*. The top panel displays the infections that occurred during the quarantine period in Sydney on board the ship. The bottom panel compares the total infections and deaths that occurred with respect to the initial population size by group.

Table S2: Number of individuals who were removed from the *Medic* after arriving at the Quarantine Station and their disease status.

| Day | Healthy removals | Total infectious/infected Removals | Mild cases | Severe cases | Unknown (mild/severe) cases |
|---|---|---|---|---|---|
| 20 | 0 | 82 | 50 | 32 | 0 |
| 21 | 0 | 57 | 43 | 14 | 0 |
| 22 | 34 | 0 | 0 | 0 | 0 |
| 23 | 151 | 0 | 0 | 0 | 0 |
| 24 | 0 | 38 | 0 | 38 | 0 |
| 25 | 0 | 108 | 0 | 0 | 108 |

#### S2 Model formulation

##### S2.1 Model state and parameter description

The states and parameters that are used in Figure 2 of the main manuscript are described below:

State description for Group $i$:

1. $\mathbf{S}_i$ **:** Completely susceptible state.
2. $\mathbf{E}_i$ **:** Exposed state.
3. $\mathbf{A}_i$ **:** Asymptomatic state.
4. $\mathbf{R}_{Ai}$ **:** Recovered state of asymptomatic individuals.
5. $\mathbf{I}_{iP}$ **:** Pre-symptomatic state.
6. $\mathbf{I}_{iS}$ **:** Symptomatic severe influenza.
7. $\mathbf{M}_i$ **:** Symptomatic mild influenza.
8. $\mathbf{C}_i$ **:** Recovering individuals with severe influenza symptoms. They are no longer infectious.
9. $\mathbf{C}_{Mi}$ **:** Recovering individuals with mild influenza symptoms. They are no longer infectious.
10. $\mathbf{R}_i$ **:** Recovered state of symptomatic individuals.
11. $\mathbf{I}_{iQ}$ **:** Infectious individuals who are at the Quarantine Station.
12. $\mathbf{C}_{iQ}$ **:** Recovering individuals in the Quarantine Station. They are no longer infectious.
13. $\mathbf{R}_{iQ}$ **:** Recovered individuals at the Quarantine Station.

Parameter description for Group i:

1. $\beta_{ijI}$ : Transmission rate from Group $j$ to $i$ of symptomatic influenza cases.
2. $\beta_{ijA}$ : Transmission rate from Group $j$ to $i$ of asymptomatic influenza cases.
3. $\delta_i(1-\omega_i)$ : Rate of progression from exposed to asymptomatic infectious states.
4. $\gamma_i$ : The recovery rate of infectious asymptomatic individuals. Also the rate of progression of symptomatic infectious to recovering state.
5. $\delta_i\omega_i$ : Rate of progression from exposed to pre-symptomatic infectious states.
6. $\alpha_i(1-\tau_i)$: Rate of progression of from pre-symptomatic infectious to symptomatic severe infectious states.
7. $\alpha_i\tau_i$: Rate of progression from pre-symptomatic infectious to symptomatic mild infectious states.
8. $\gamma_{iJ}$ : Rate of progression from symptomatic infectious to isolation state.
9. $\gamma_{Hi}$: Rate of progression s from symptomatic infectious to hospitalisation state.
10. $\nu_i$: The progression rate from recovery to fully recovered state.
11. $\epsilon_i(t)$ : Rate of removal of healthy people.
12. $\zeta_i(t)$: Rate of removal of cases (infectious or infected).
13. $d_i$: Death rate of individuals due to influenza.

##### S2.2 Force of infection

The force of infection, $\lambda_i(t)$, acting on Group $i$ at time $t$ is

$$\lambda_i(t) = \sum_{j=1}^{2} \frac{(\beta_{ijA} A_j(t) + \beta_{ijI} I_j(t))}{N_j(t)},$$

where

$$I_j(t) = I_{jP}(t) + I_{jS}(t) + M_j(t)$$

and $N_j(t)$ is the size of the $j$th group at time $t$ and $N(t) = \sum_{j=1}^{2} N_j(t)$.

##### S2.3 Time dependent parameters

Rate of removal of healthy people ($\epsilon_i(t)$) and the rate of removal of cases ($\zeta_i(t)$) are taken as time-dependent parameters.

#### S3 Removal of healthy, infectious and infected people

We will take the rates of removal of healthy ($\epsilon_i(t)$), infectious and infected people ($\zeta_i(t)$) as time-dependent across the groups. These parameters will need to be estimated except when it is reasonable to assume the rates are zero. For example, for $t_0, t_a$ discrete time (i.e., days) period, if the ship record has no mention of the removal of people within the time period, $\epsilon_i(t)$ and $\zeta_i(t)$ can be considered as zero.

If the ship record mentions the removal of healthy people on the day $t_b$, and removal of infectious infected people on the day $t_c$, the rates of removal of healthy people and infectious infected people in Group $i$, $\epsilon_i(t_b)$ and $\zeta_i(t_c)$ will be estimated as part of the parameter estimation.

Table S3: Rates of removal of healthy, infectious and infected people for $i$th groups

| | Rates(per day) | | |
|---|---|---|---|
| **Time period** | **Removal of healthy** | **Removal of infectious/ infected** | |
| $0 - t_a$ | 0 | 0 | No removals |
| $t_b$ | $\epsilon_i(t_b)$ | 0 | Removal of healthy |
| $t_c$ | 0 | $\zeta_i(t_c)$ | Removal of infectious |

#### S4 Model assumptions

**Assumption 1:** Rates of transmission between asymptomatic ($\beta_{ijA}$) and symptomatic infectious ($\beta_{ijI}$) are the same and will be denoted as $\beta_{ij}$.

**Reason:** Although research shows that for influenza, transmission between asymptomatic individuals may be less than that between symptomatic individuals, evidence regarding the 1918-19 strain is insufficient. To reduce model complexity we will be making this assumption.

**Assumption 2:** Except for transmission parameters $\beta_{ij}$, all the other parameters across the groups will be assumed to be equal.

**Reason:** Parameters of diseases related factors such as incubation period, pre-symptomatic period, infectious period, recovering period, and recovery period will be similar across crew, troops and civilians as they are generally exposed to a single influenza strain in the ship. By making this assumption, instead of having to infer, at most, three sets of disease-related parameter sets except for $\beta_{ij}$, we will only be estimating $\beta_{ij}$ and one set of other parameters. Hence, the number of parameters that need to be estimated becomes less.

**Assumption 3:** Number of groups with different mixing behaviour can be minimised to two instead of three.

**Reason:** The majority of the ships' populations included crew and troops only. *Medic* only had four civilians on board. *Devon* had a significant number of civilians and the number was 66. Therefore, it is reasonable to take that all the ships had two groups on board. By considering two groups (crew and passengers) instead of three, the number of $\beta_{ij}$ parameters that need to be estimated becomes 4 instead of 9.

**Assumption 4:** Time-dependent parameters, rates of removal of healthy and infectious/ infected for a particular day are the same across all the groups.

**Reason:** This assumption is made to reduce the number of parameters that need to be estimated. As the number of time-dependent parameters that need to be estimated increases with the number of days where population fluctuations take place, this will add to the number of total parameters that need to be estimated. Therefore, to reduce the dimensionality of the parameter space that needs to be estimated, we will make this assumption.

**Assumption 5:** Initial conditions of the ship outbreaks will be assumed to be known.

**Reason:** The ship outbreak details are generally well-recorded and sufficient to make reliable assumptions with regard to the initial conditions of the outbreak.

##### S4.1 Initial conditions of each ship

We assume that the initial conditions of the ships are known and this information was gathered from the available data.

***Medic***: Not clear which group started the epidemic. We start the epidemic by randomly choosing an individual from either crew or troops and assumed that that person was exposed to influenza by the time ship left the source of infection.

***Boonah***: One of the troops starts the epidemic. We assume that the soldier was in the exposed state when the ship left the source of infection.

***Devon***: Assume that one exposed troop started the epidemic by the time the ship left the source of infection.

***Manuka***: Assume that one crew started the epidemic and this person was exposed by the time ship left the source of infection.

##### S4.2 Sample path generation

We generate exact sample paths using the Doob-Gillespie (Gillespie, 1977; Doob, 1945) algorithm with changes to address the population size fluctuations that occur due to removal of healthy, infectious or infected individuals.

Let the initial state of the chosen stochastic model be denoted from the vector $\boldsymbol{U(t_0)}$ where $t_0$ is the time when the ship arrived at the source of infection or else, the time at which the epidemic is assumed to have started. Consider $K$ groups. Let the chosen fixed (i.e., not time-dependent) set of parameters be denoted as $\mathbf{\Theta}$ and time-dependent parameters between $t_0$ to $t_a$, the time where the next population fluctuation occurs, be denoted as $\epsilon_i(t_0)$ and $\zeta_i(t_0)$.

We will use the following methodology to generate sample paths.

1. Given the parameters $\mathbf{\Theta}$ and $\epsilon_i(t_0)$ and $\zeta_i(t_0)$, generate a Gillespie sample path from time $t = t_0$ to $t = t_a$ where the first fluctuation of the ship's population occurred. At time $t_a$, let the state of the Gillespie path be denoted as $\mathbf{U}(t_a)$.

2. On day $t = t_a$, we will take that a total of $n(t_a)$ people left the ship. If the change in population size occurred as
   (a) **Removal of healthy, infectious or infected people:**
   Consider the time period $t = t_a - t_b$, where $t_b$ is the day when the next population fluctuation takes place. Change time-dependent parameters accordingly, say $\epsilon_i(t_a)$ and $\zeta_i(t_a)$. Generate a Gillespie sample path from time $t = t_a$ to $t = t_b$ using parameters $\mathbf{\Theta}$, $\epsilon_i(t_a)$ and $\zeta_i(t_a)$ and initial condition $\mathbf{U}(t_a)$.
   At time $t_b$, the state of the sample path is $\boldsymbol{U}(t_b)$

3. Repeat Step 2 until the sample path is completed at $t = T$.

**Note:** See Supplementary Material 2 for the details of our parameter estimation framework.

#### S5 Studying the effects of quarantine measures

To study the effects of quarantine measures, we considered outbreaks of *Medic* and *Boonah* using conditional re-sampled and re-sampled paths.

To generate re-sampled paths, for each estimated parameter set, we generated sample paths with and without interventions. We made sure that the sample paths under both models up to arriving at a Quarantine Station were identical by controlling the random number generation in Matlab.

##### S5.1 Conditional re-sampled and re-sampled paths of *Devon*

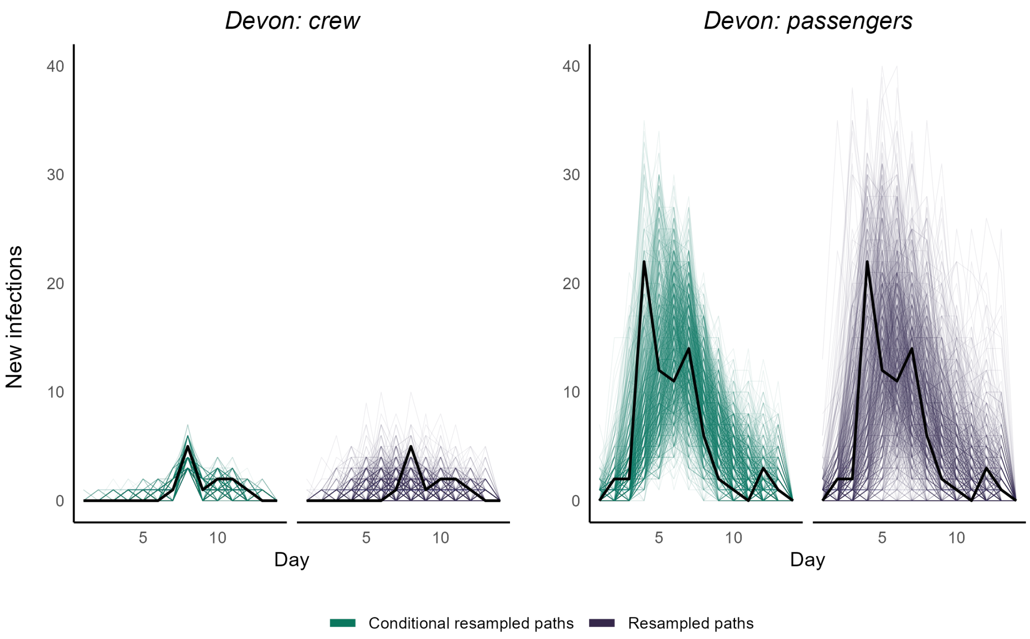


Figure S4: Conditional re-sampled paths (in green) by the hierarchical estimation algorithm and re-sampled paths (in purple) by the estimated parameters. Black lines are the observed data.

##### S5.2 *Medic*

Table S4: Difference between no interventions and interventions (total infections throughout the epidemic) for *Medic*

| | Within passengers | | | Within crew | | |
|---|---|---|---|---|---|---|
| | Median (25, 75)% | % increase Median (25, 75)% | Proportion of positive difference | Median (25, 75)% | % increase Median (25, 75)% | Proportion of positive difference |
| **Conditional** | 17 (5, 29) | 7.52 (2.13, 12.92) | 0.822 | 1 (-3, 5) | 2.24(-6.38, 11.90) | 0.409 |
| **Re-sampled** | 11 (2, 22) | 5.43(1.97, 10.42) | 0.783 | 0 (-3, 3) | 0 (-7.14, 7.45) | 0.407 |

##### S5.2.1 Crew

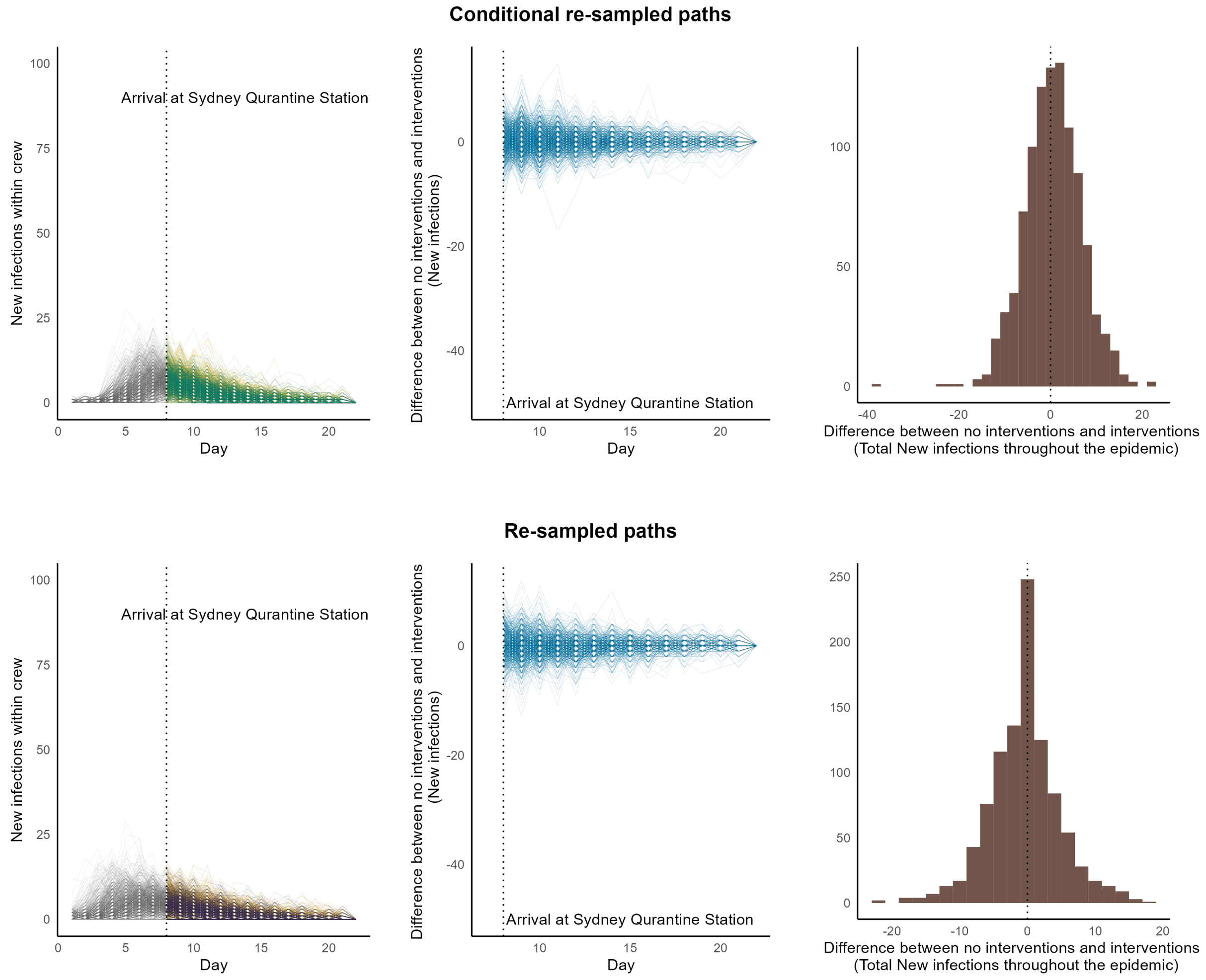


Figure S5: Interventions vs. no interventions for crew in *Medic.*
**Simulated paths in Grey**: Trajectories up to the time the ship arrived at the quarantine Station. **Simulated paths in Green:** Conditional re-sampled paths. **Simulated paths in purple:** re-sampled paths. **Simulated paths in yellow:** Counterfactual paths of no interventions corresponding to conditional re-sampled or re-sampled paths.

##### S5.2.2 Passengers

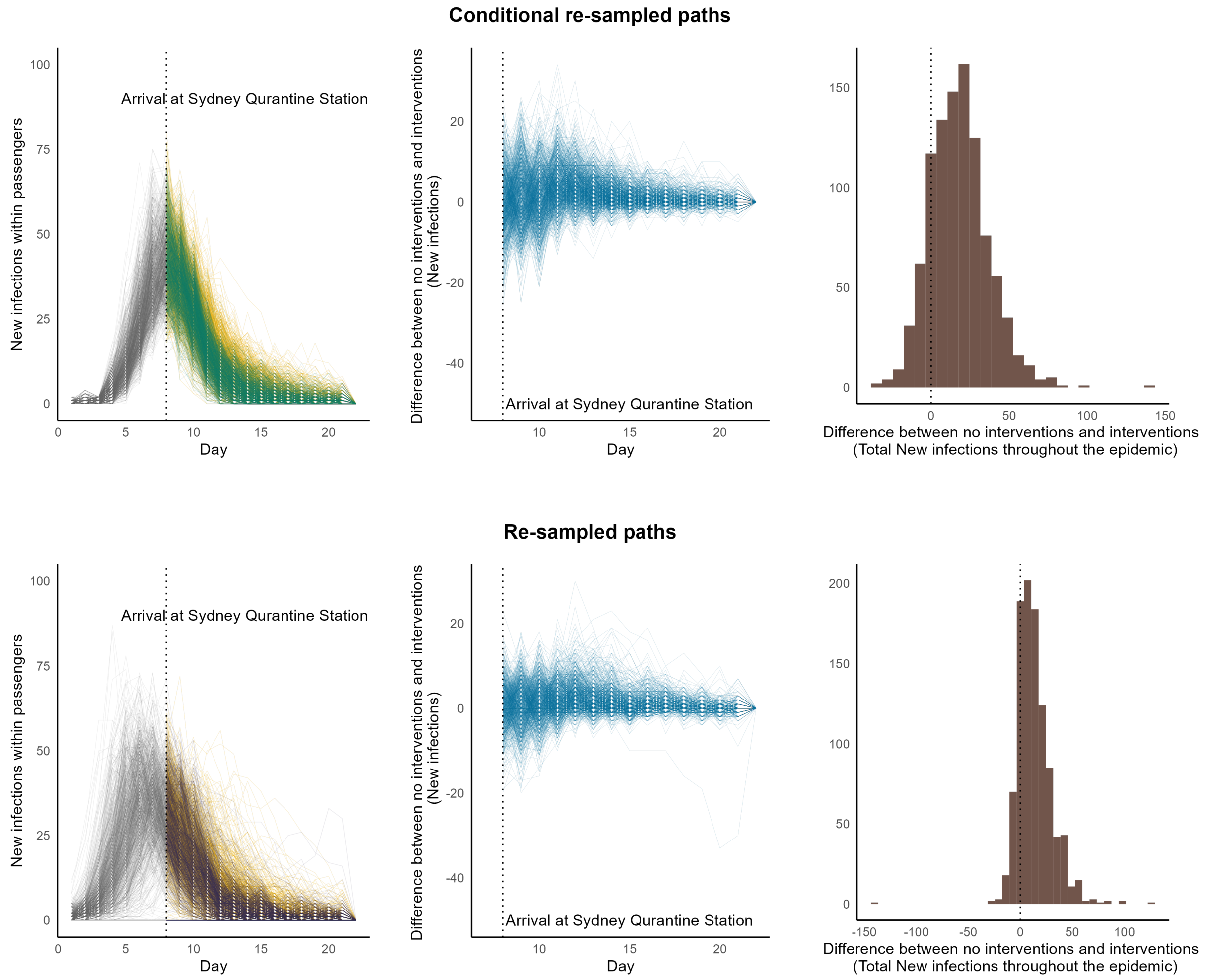


Figure S6: Interventions vs. no interventions for passengers in *Medic.*
**Simulated paths in Grey**: Trajectories up to the time the ship arrived at the quarantine Station. **Simulated paths in Green:** Conditional re-sampled paths. **Simulated paths in purple:** re-sampled paths. **Simulated paths in yellow:** Counterfactual paths of no interventions corresponding to conditional re-sampled or re-sampled paths.

##### S5.2.3 Deaths

Table S5: Difference between no interventions and interventions (total deaths throughout the epidemic) for *Medic*

| | Median (25, 75)% | % increase Median (25, 75)% | Proportion of positive difference |
|---|---|---|---|
| **Conditional** | 2 (-2, 7) | 9.52 (-9.76, 31.25) | 0.592 |
| **Re-sampled** | 0 (-3, 3) | 0 (-400, 600) | 0.45 |

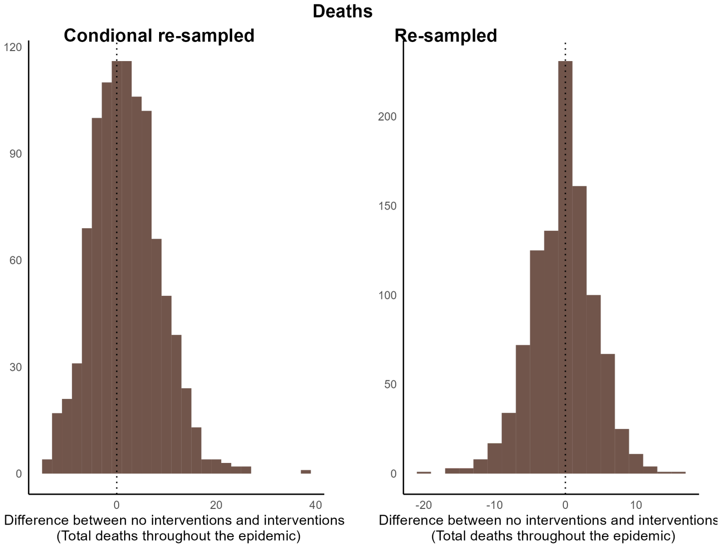


Figure S7: Interventions vs. no interventions for passengers in *Manuka*.

##### S5.3 *Boonah*

###### S5.3.1 Crew

Table S6: Difference between no interventions and interventions (total infections throughout the epidemic) for *Boonah*

| | **Median (25, 75)%** | **% increase Median (25, 75)%** | **Proportion of positive difference** |
|---|---|---|---|
| **Conditional** | 0 (0, 1) | 0 (0, 3.25) | 0.401 |
| **Re-sampled** | 0 (0, 0) | 0 (0, 0) | 0.034 |

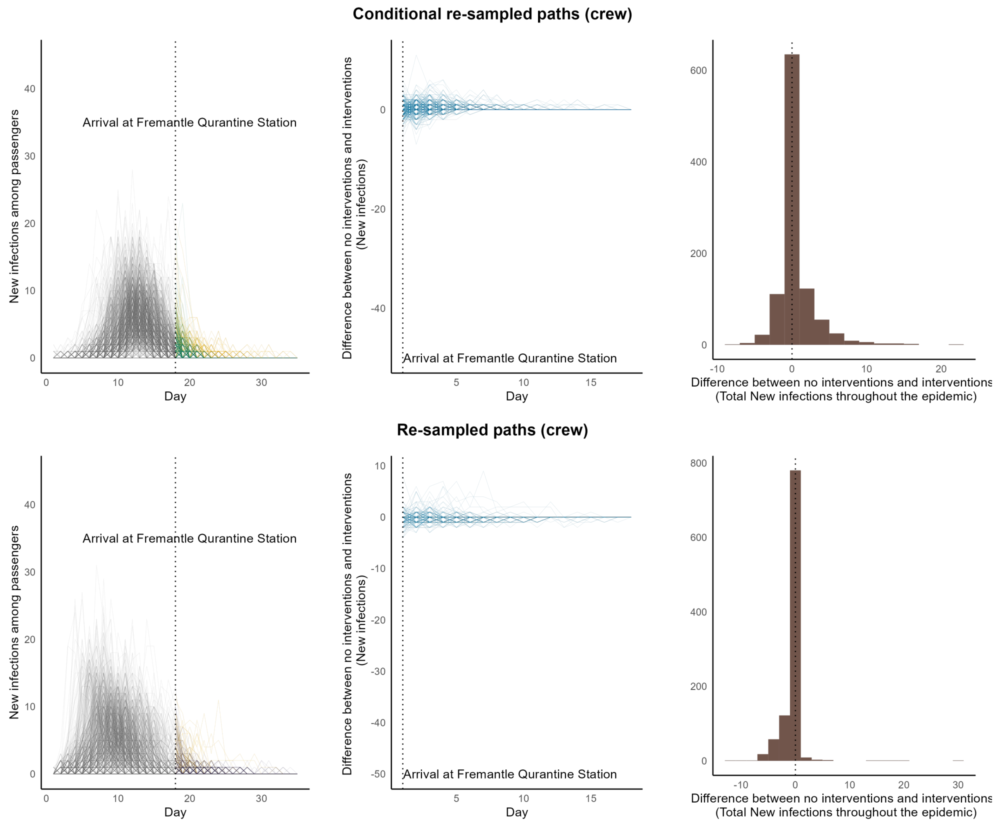


Figure S8: Interventions vs. no interventions for crew in *Boonah.*
**Simulated paths in Grey**: Trajectories up to the time the ship arrived at the quarantine Station. **Simulated paths in Green:** Conditional re-sampled paths. **Simulated paths in purple:** re-sampled paths. **Simulated paths in yellow:** Counterfactual paths of no interventions corresponding to conditional re-sampled or re-sampled paths.

##### S5.3.2 Deaths

Table S7: Difference between no interventions and interventions (total deaths throughout the epidemic) for *Boonah*

| | Median (25, 75)% | % increase Median (25, 75)% | Proportion of positive difference |
|---|---|---|---|
| **Conditional** | 0 (-2, 2) | 0 | 0.451 |
| **Re-sampled** | 0 (-4, 0) | 0 | 0.107 |

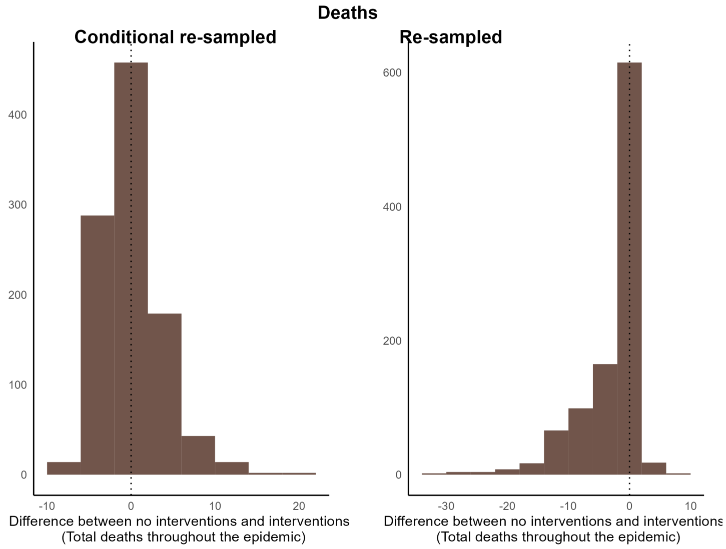


Figure S9: Interventions vs. no interventions of deaths in *Boonah*.

#### S6 Calculation of $R_0$

Infectious classes: $E_1, E_2, A_{11}, A_{12}, A_{21}, A_{22}, I_{1p}, I_{2p}, M_1, M_2, I_{1s}, I_{2s}$

$$F=\begin{pmatrix}0&0&\beta_{11}&\beta_{12}&\beta_{11}&\beta_{12}&\beta_{11}&\beta_{12}&\beta_{11}&\beta_{12}&\beta_{11}&\beta_{12}\\0&0&\beta_{21}&\beta_{22}&\beta_{21}&\beta_{22}&\beta_{21}&\beta_{22}&\beta_{21}&\beta_{22}&\beta_{21}&\beta_{22}\\0&0&0&0&0&0&0&0&0&0&0&0\\0&0&0&0&0&0&0&0&0&0&0&0\\0&0&0&0&0&0&0&0&0&0&0&0\\0&0&0&0&0&0&0&0&0&0&0&0\\0&0&0&0&0&0&0&0&0&0&0&0\\0&0&0&0&0&0&0&0&0&0&0&0\\0&0&0&0&0&0&0&0&0&0&0&0\\0&0&0&0&0&0&0&0&0&0&0&0\\0&0&0&0&0&0&0&0&0&0&0&0\\0&0&0&0&0&0&0&0&0&0&0&0\end{pmatrix}$$

$$V=\begin{pmatrix}\delta&0&0&0&0&0&0&0&0&0&0&0\\0&\delta&0&0&0&0&0&0&0&0&0&0\\-\delta*(1-\omega)&0&\alpha&0&0&0&0&0&0&0&0&0\\0&-\delta*(1-\omega)&0&\alpha&0&0&0&0&0&0&0&0\\0&0&-\alpha&0&\gamma&0&0&0&0&0&0&0\\0&0&0&\alpha&0&\gamma&0&0&0&0&0&0\\-\delta*\omega&0&0&0&0&0&\alpha&0&0&0&0&0\\0&-\delta*\omega&0&0&0&0&0&\alpha&0&0&0&0\\0&0&0&0&0&0&-\alpha*\tau&0&\gamma&0&0&0\\0&0&0&0&0&0&0&-\alpha*\tau&0&\gamma&0&0\\0&0&0&0&0&0&-\alpha*(1-\tau)&0&0&0&(\gamma+d_1)&0\\0&0&0&0&0&0&0&-\alpha*(1-\tau)&0&0&0&(\gamma+d_2)\end{pmatrix}$$
