## Supplementary material 2 for "How effective were Australian Quarantine Stations in mitigating transmission aboard ships during the influenza pandemic of 1918-19?"

### How effective were Australian Quarantine Stations in mitigating transmission aboard ships during the influenza pandemic of 1918-19? Supplementary Material 2 Parameter estimation

Punya Alahakoon[1,3,4], Peter G. Taylor[1], James M. McCaw[*,2]

[1]School of Mathematics and Statistics , The University of Melbourne, Melbourne, Australia.
[2]Centre for Epidemiology and Biostatistics, Melbourne School of Population and Global Health, The University of Melbourne, Melbourne, Australia.
[3]School of Population Health, University of New South Wales, Sydney, Australia.
[4]Kirby Institute, University of New South Wales, Sydney, Australia.

#### S1 Introduction

In this section, we explain in detail how the parameter estimation was carried out, particularly for the outbreak onboard *Medic*. Details of other ships, if not included in this document, are included in GitHub (details are provided at the bottom of this document).

We used the two-step algorithm of Alahakoon, McCaw, and Taylor (2022) to estimate the model parameters within a hierarchical framework. This method uses Approximate Bayesian Computation (ABC) and pseudo-marginal methods. The first step of the algorithm considers each outbreak independently and estimates the parameters. We used the ABC-SMC algorithm of Toni, Welch, Strelkowa, Ipsen, and Stumpf (2009) for this purpose and the details are described below. We used normal densities with standard deviations estimated from the particles from the previous generations as the perturbation kernel when the ABC-SMC algorithm was run for each outbreak independently. Prior to conducting parameter estimation, we identified the non-implausible parameter spaces and this is also described below for *Medic*. Suitable tolerance values were chosen by running basic ABC algorithms with high tolerance values and displaying the sample of parameter values and distance metrics (similar to the approach described in Alahakoon et al. (2022)).

Then we estimated the hyper-parameters of the outbreaks under a hierarchical framework. We assumed that only the $\beta_{ij}$ are sampled from a common probability distribution. We assume that this distribution, that is the conditional prior distribution, is a truncated multivariate normal distribution. We denote $\Psi_{\beta_{ij}}$ as the hyper-mean of the transmission from group $j$ to $i$, $\sigma_{\beta_{ij}}$ as the standard deviation hyper-parameter from group $j$ to $i$.

The details of the prior distributions and the conditional prior distributions are:

$$\begin{aligned}
\Psi_{\beta_{11}} &\sim \text{Uniform}(0.001, 10) \\
\Psi_{\beta_{12}} &\sim \text{Uniform}(0.001, 10) \\
\Psi_{\beta_{22}} &\sim \text{Uniform}(0.001, 6) \\
\Psi_{\beta_{21}} &\sim \text{Uniform}(0.001, 5) \\
\sigma_{\beta_{11}}, \sigma_{\beta_{12}}, \sigma_{\beta_{22}} &\sim \text{Uniform}(0, 3.5) \\
\sigma_{\beta_{21}} &\sim \text{Uniform}(0, 2.5) \\
R \sim \text{LKJcorr}(2) &\quad \text{(Prior for correlation matrix)} \\
\beta_{ij} \sim \text{Truncated Multivariate Normal}&(\mathbf{\Psi}, \Sigma, \mathbf{a}, \mathbf{b}),
\end{aligned}$$

*

where,

$$R = \begin{bmatrix} 1 & \rho_{12} & \rho_{13} & \rho_{14} \\ \rho_{21} & 1 & \rho_{23} & \rho_{24} \\ \rho_{31} & \rho_{32} & 1 & \rho_{34} \\ \rho_{41} & \rho_{42} & \rho_{43} & 1 \end{bmatrix} \tag{S.1}$$

the covariance matrix

$$\Sigma = \begin{bmatrix} \sigma_{\beta_{11}} & 0 & 0 & 0 \\ & \sigma_{\beta_{22}} & 0 & 0 \\ 0 & 0 & \sigma_{\beta_{33}} & 0 \\ 0 & 0 & 0 & \sigma_{\beta_{44}} \end{bmatrix} R \begin{bmatrix} \sigma_{\beta_{11}} & 0 & 0 & 0 \\ & \sigma_{\beta_{22}} & 0 & 0 \\ 0 & 0 & \sigma_{\beta_{33}} & 0 \\ 0 & 0 & 0 & \sigma_{\beta_{44}} \end{bmatrix} \tag{S.2}$$

and $\mathbf{a} = (0.001, 0.001, 0.001, 0.001)$ and $\mathbf{b} = (10, 10, 6, 5)$. We used a pseudo-marginal framework to estimate the hyperparameters. The details are explained in this document.

Once hyperparameters were estimated, we then estimated the ship-specific parameters using an ABC algorithm. Here, we proposed the transmission rates using the conditional prior, $TruncatedMultivariateNormal(\mathbf{\Psi}, \Sigma, \mathbf{a}, \mathbf{b})$, with the estimated hyper-parameters. All the other parameters were sampled by perturbing from the last generation of the marginal posterior distributions in Step 1(a). In this step, we used the final generations' tolerance values of the ABC-SMC algorithm we used in the first step.

#### S2 Identifying the non-implausible regions of parameter spaces

Before conducting parameter estimation, we identified the plausible regions of the parameter space for the outbreak in *Medic* from the following procedure:

1. Initially we considered distributions to generate parameters that needed to be estimated as follows:

Table S1

| Parameter | Distribution |
|---|---|
| $\beta_{ij}$ for $i, j = 1, 2$ | uniform(0.001,10) |
| $1/\delta(1-\omega)$ | lognormal (0.4039,0.6) |
| $1/\delta\omega$ | lognormal (0.4039,0.25) |
| $\tau$ | uniform(0.0001,1) |
| $1/\alpha(1-\tau)$ | lognormal (-0.2342,0.35) |
| $1/\nu$ | lognormal(1.8318,0.6) |
| $d_{Crew}$ | beta (1.5,30) |
| $d_{Passengers}$ | beta (1.5,30) |
| $\epsilon(10), \epsilon(11), \zeta(8), \zeta(9), \zeta(12), \zeta(14)$ | uniform(0.001,10) |

   When identifying plausible parameter spaces for the parameters, we only intend to identify the parameter regions of $\beta_{ij}$ for $i, j = 1, 2$, $\epsilon(10), \epsilon(11), \zeta(8), \zeta(9), \zeta(12)$, and $\zeta(14)$. During the parameter estimation stage for these parameters, we intend to use uniform prior distributions in pre-specified regions identified at this stage. For all the other parameters, we intend to use informative prior distributions and therefore, plausible regions for these will not be identified.

2. Then, using a Latin Square Design, we generated parameter combinations (number of parameters $\times 75 = 18 \times 75 = 1350$ for all the above parameters. we simulated data (from the Model as in Figure 3) 50 times under each parameter combination. From these data, we calculated the average of the summary statistics (Euclidean distance, absolute difference between generated and observed data) we plan to use in the parameter estimation stage (ABC algorithm) to check which parameter combinations generated expected results. See Figure 4. This step is called Wave 1.

3. From the above step, we identify the plausible regions for $\beta_{ij}$s and for $\zeta(8)$. And then we change the intervals of the uniform distributions accordingly in which the Latin Square was generated.

4. Repeat Step 2 and denote it as Wave 2. Repeat Step 3 until reasonably plausible parameter spaces are identified for $\beta_{ij}$ for $i, j = 1, 2$, $\epsilon(10), \epsilon(11), \zeta(8), \zeta(9), \zeta(12)$, and $\zeta(14)$. See the Figures below.

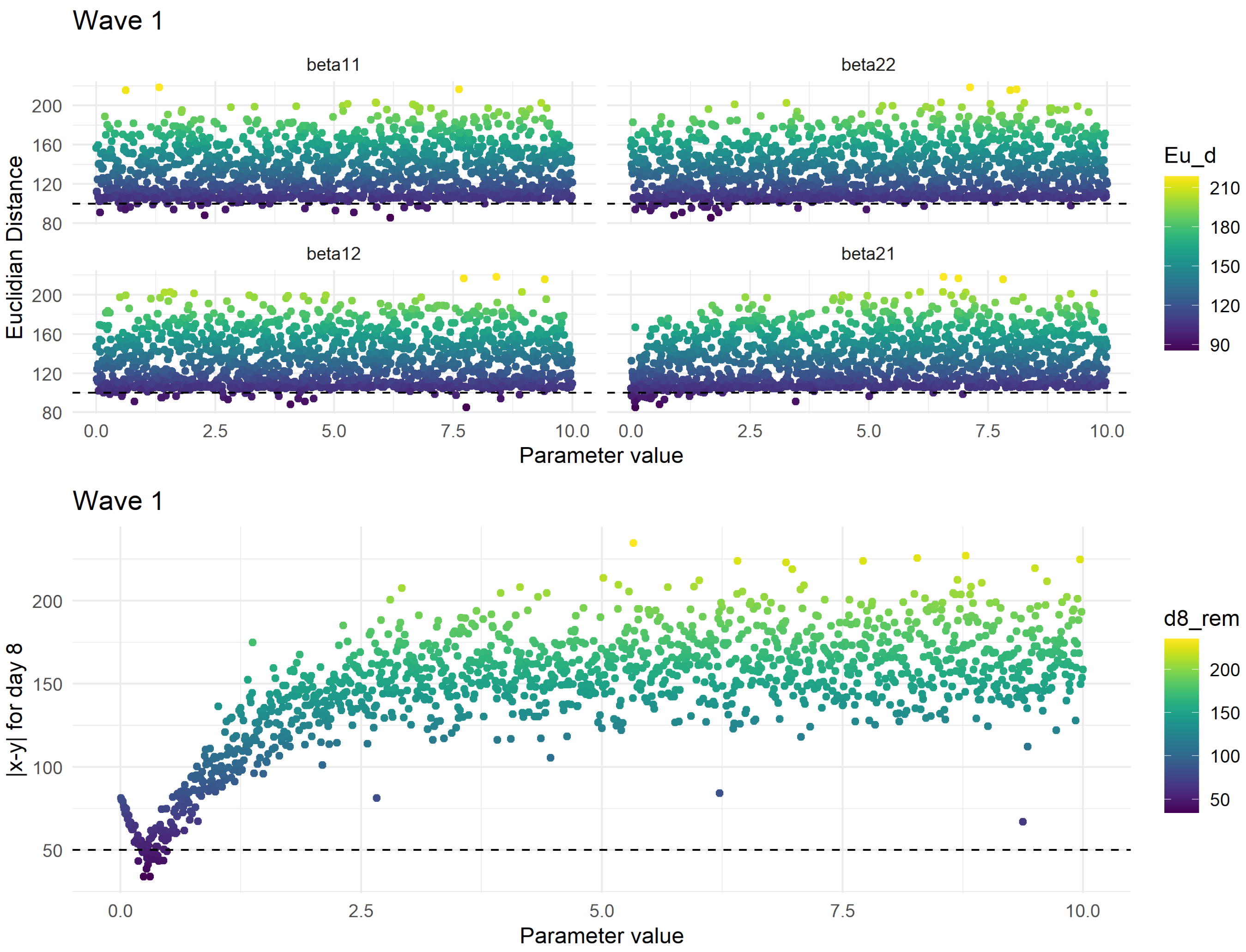


Figure S1: Wave 1

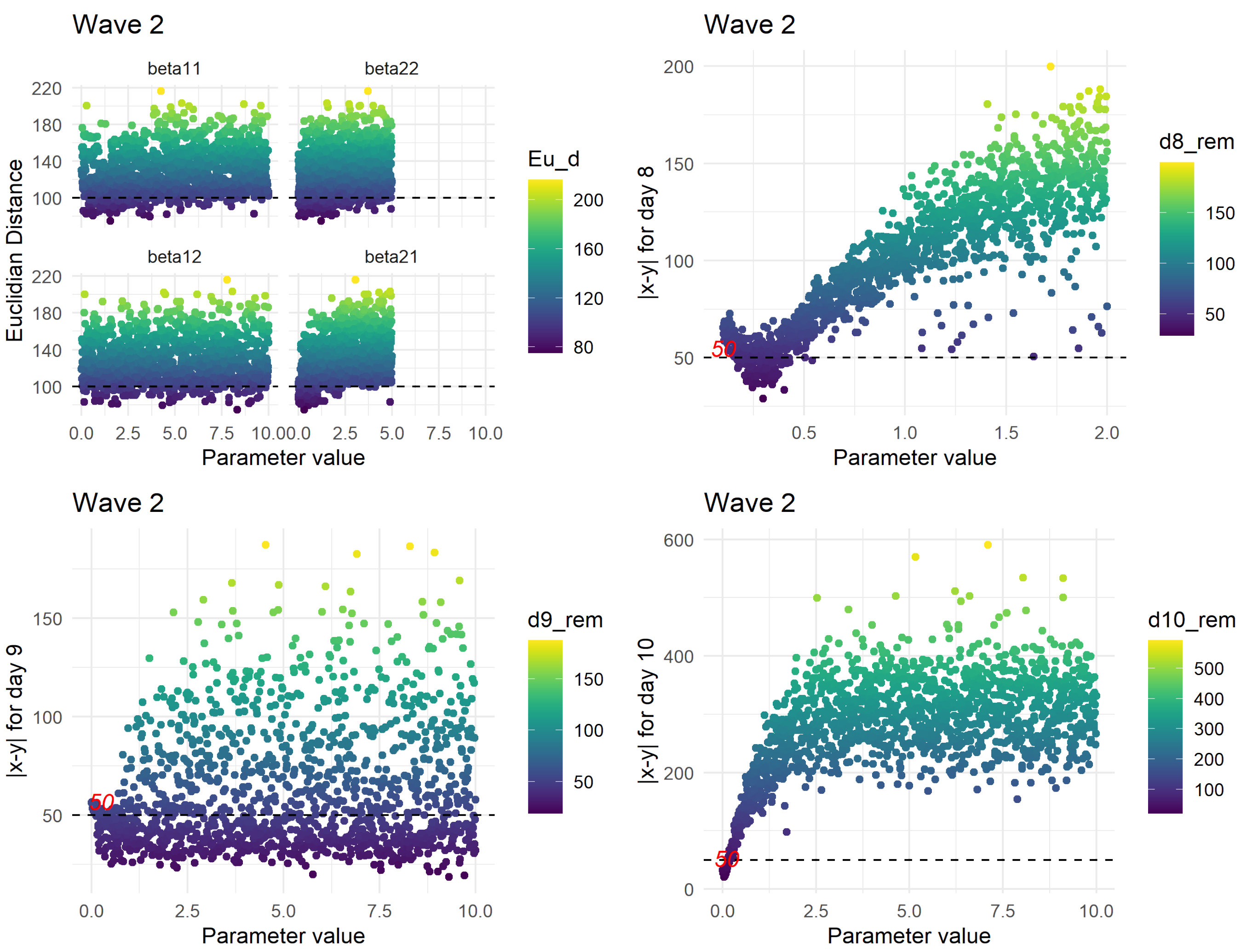


Figure S2: Wave 2

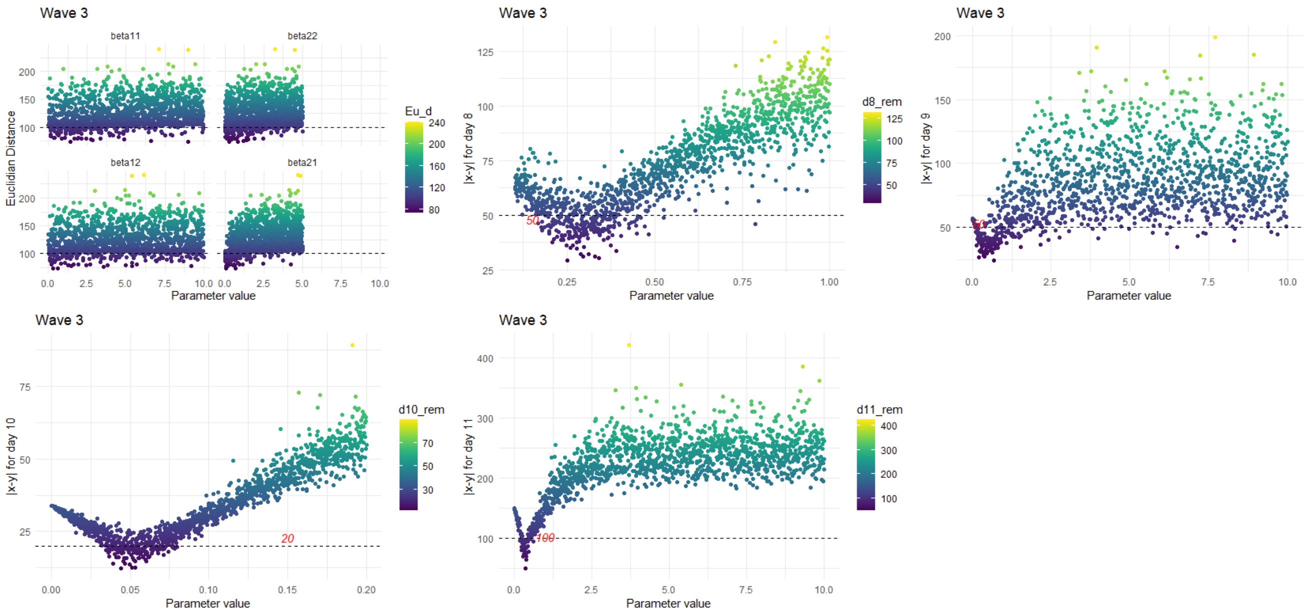


Figure S3: Wave 3

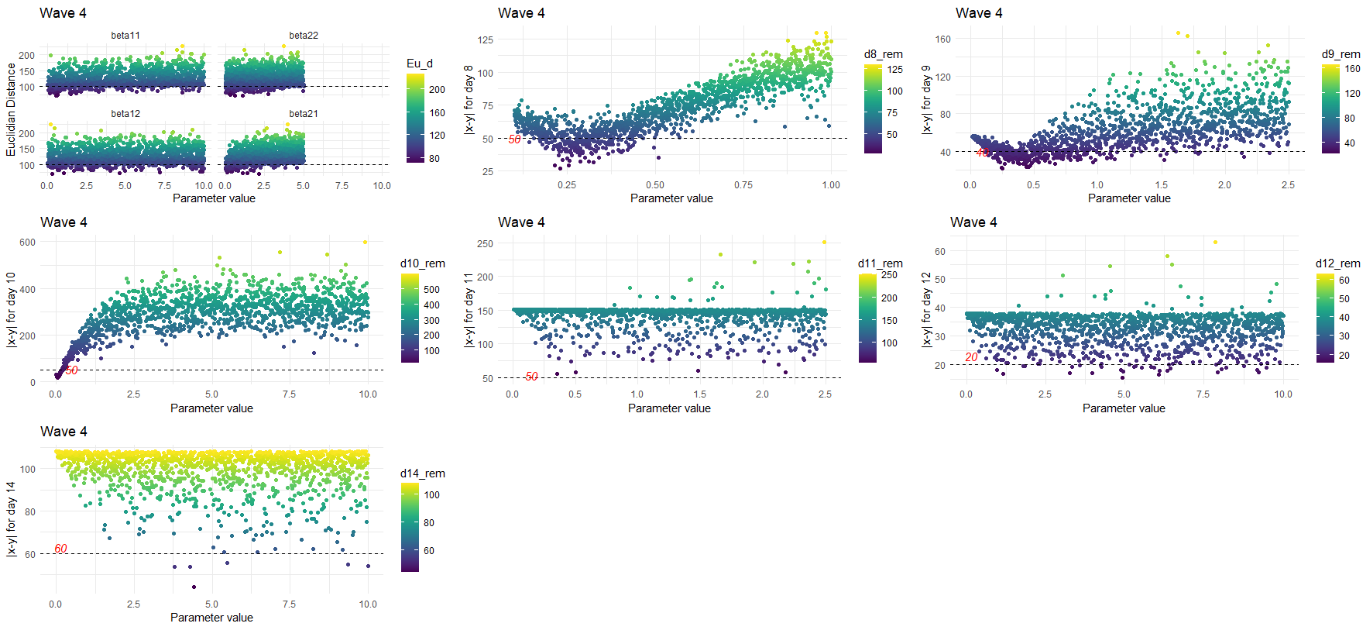


Figure S4: Wave 4

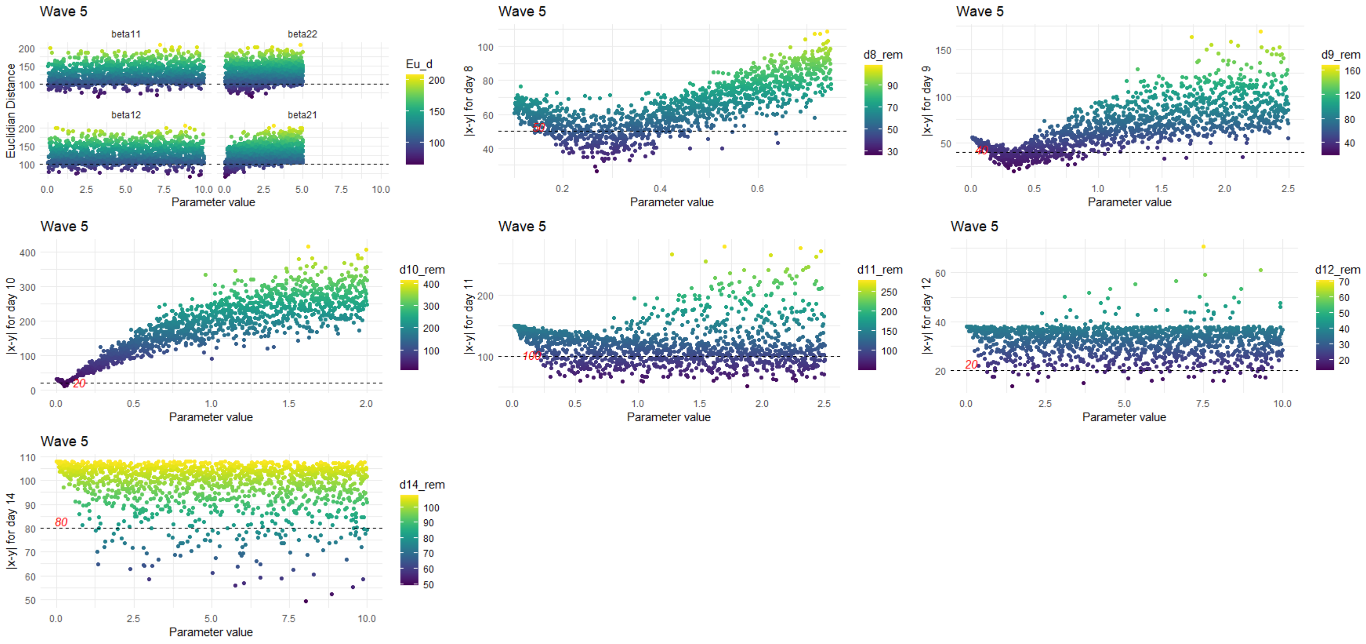


Figure S5: Wave 5

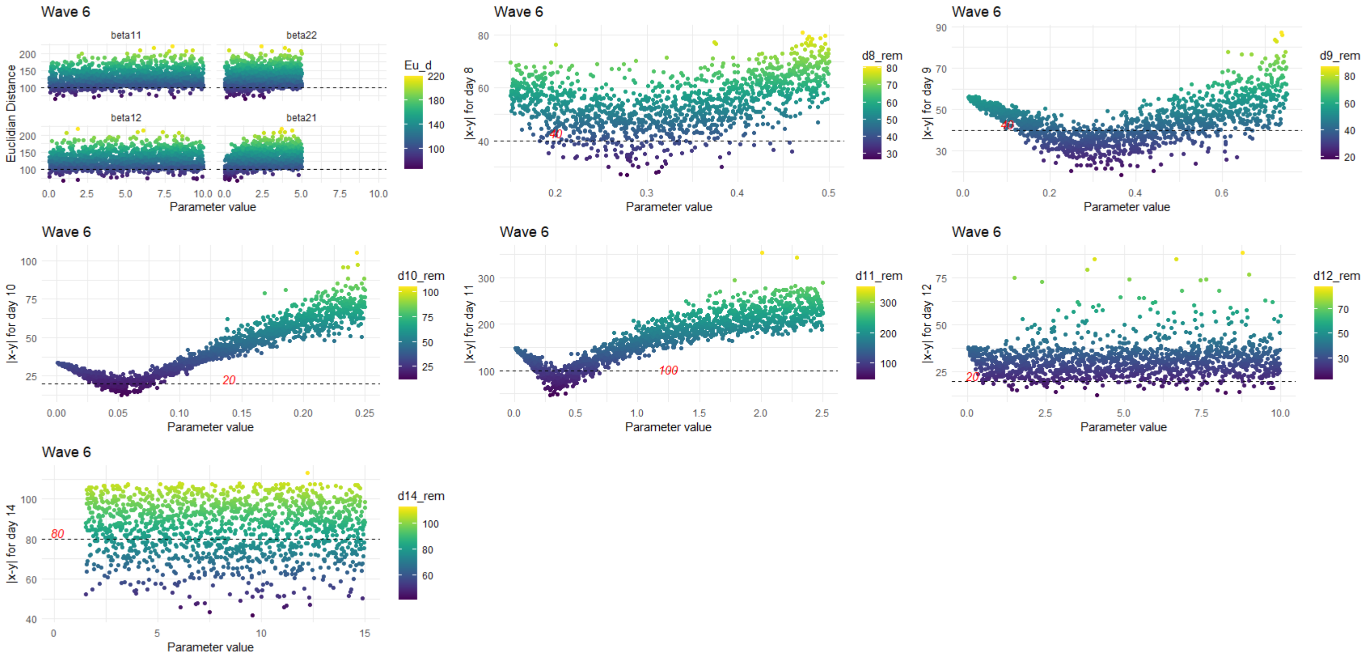


Figure S6: Wave 6

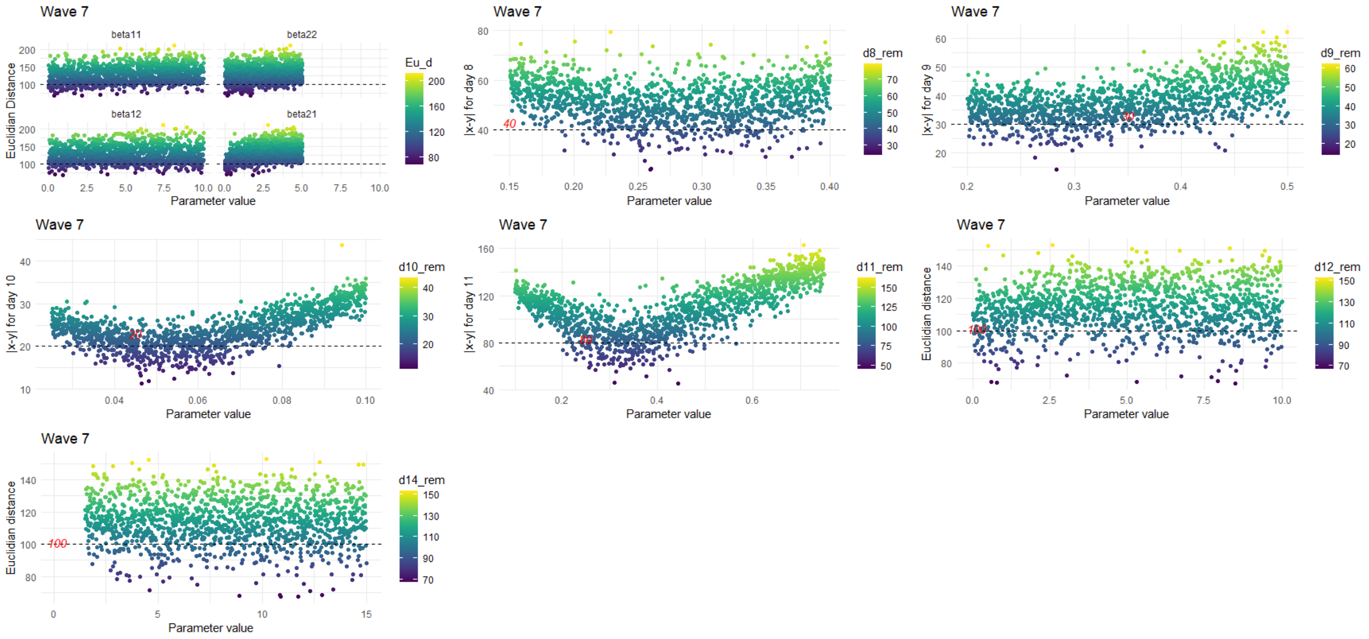


Figure S7: Wave 7

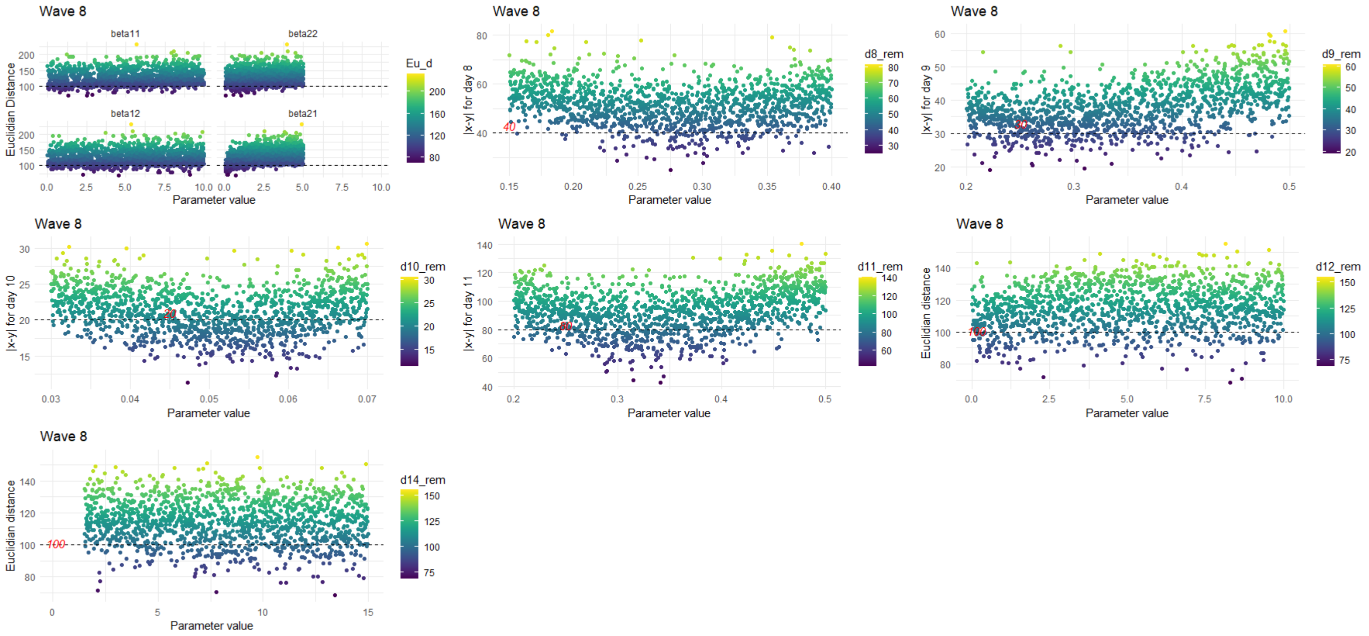


Figure S8: Wave 8

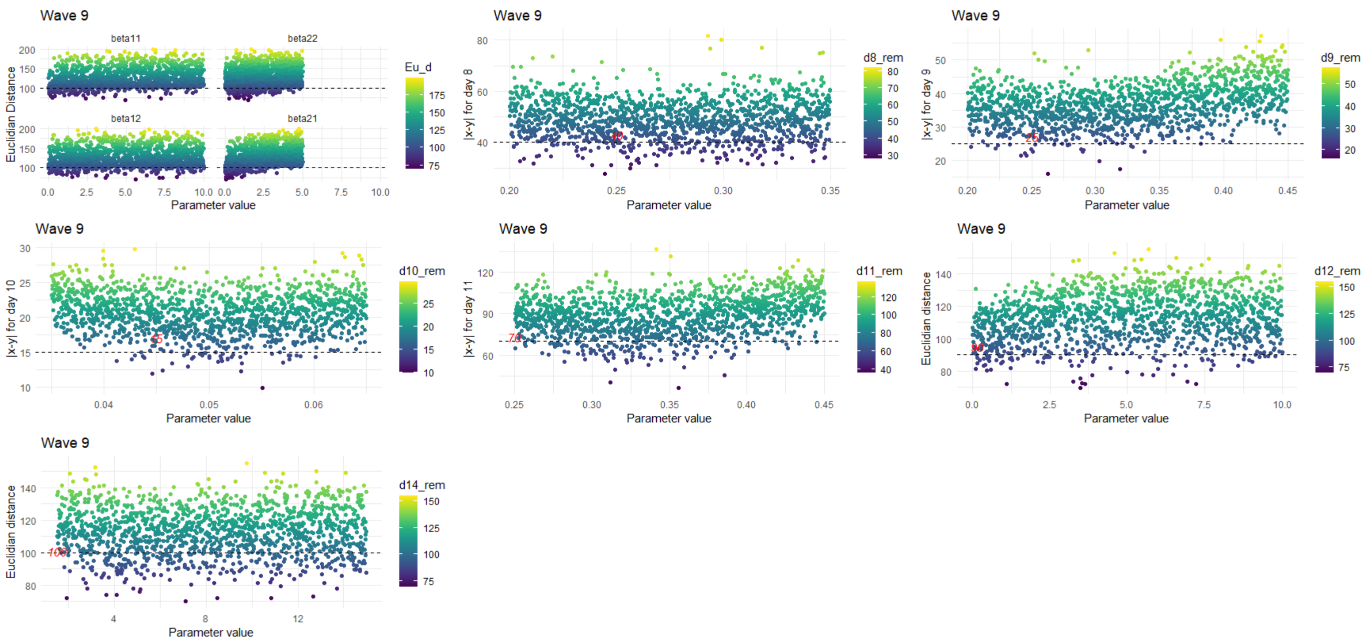


Figure S9: Wave 9

In this step, we could not find plausible regions for $\beta_{11}, \beta_{12}$, and $\zeta(14)$.

**Please refer to GitHub files (link provided below) for details on finding the non-implausible regions of other ships.**

### S3 Parameter Estimation using the ABC-SMC algorithm

#### S3.1 Prior distributions

After identifying the plausible regions for parameters from the previous step, we used the following distributions as prior distributions for the parameters:

Table S2: Prior distributions for *Medic*

| Parameter | Prior distribution |
|---|---|
| $\beta_{11}$ and $\beta_{12}$ | uniform(0.001,10) |
| $\beta_{22}$ and $\beta_{21}$ | uniform(0.001,5) |
| $1/\delta(1-\omega)$ | lognormal (0.4039,0.6) |
| $1/\delta\omega$ | lognormal (0.4039,0.25) |
| $\tau$ | uniform(0.0001,1) |
| $1/\alpha(1-\tau)$ | lognormal (-0.2342,0.35) |
| $1/\nu$ | lognormal(1.8318,0.6) |
| $d_{Crew}$ | beta (1.5,30) |
| $d_{Passengers}$ | beta (1.5,30) |
| $\epsilon(10)$ | uniform(0.001,0.15) |
| $\epsilon(11)$ | uniform (0.15, 0.7) |
| $\zeta(8)$ | uniform (0.1, 2) |
| $\zeta(9)$ | uniform (0.01, 2.5) |
| $\zeta(12)$ | uniform (0.001, 10) |
| $\zeta(14)$ | uniform (1.5, 15) |

Table S3: Prior distributions for *Boonah*

| Parameter | Prior distribution |
|---|---|
| $\beta_{11}$ and $\beta_{12}$ | uniform(0.001,10) |
| $\beta_{22}$ and $\beta_{21}$ | uniform(0.001,3.5) |
| $1/\delta(1-\omega)$ | lognormal (0.4039,0.6) |
| $1/\delta\omega$ | lognormal (0.4039,0.25) |
| $\tau$ | uniform(0.0001,1) |
| $1/\alpha(1-\tau)$ | lognormal (-0.2342,0.35) |
| $1/\nu$ | lognormal(1.8318,0.6) |
| $d_{Crew}$ | beta (1.5,30) |
| $d_{Passengers}$ | beta (1.5,30) |
| $\epsilon(11)$ | uniform (0.15, 0.7) |
| $\zeta(18)$ | uniform (0.4, 2) |
| $\zeta(19)$ | uniform (0.45, 2) |
| $\zeta(20:25)$ | uniform (0.001, 10) |
| $\epsilon_{crew_{(18-26)}}$ | uniform (0.05,0.25) |
| $\epsilon_{passengers_{(18-26)}}$ | uniform (0.05,0.25) |
| $\epsilon_{passengers_{(28-29)}}$ | uniform (0, 0.5) |
| $\zeta(28)$ | uniform (0.0001, 10) |

Table S4: Prior distributions for *Devon*

| Parameter | Prior distribution |
|---|---|
| $\beta_{11}$ | uniform(0.001,5) |
| $\beta_{12}$ | uniform(0.001,3) |
| $\beta_{22}$ | uniform(0.001,6) |
| $\beta_{21}$ | uniform(0.001,3) |
| $1/\delta(1-\omega)$ | lognormal (0.4039,0.6) |
| $1/\delta\omega$ | lognormal (0.4039,0.25) |
| $\tau$ | uniform(0.0001,1) |
| $1/\alpha(1-\tau)$ | lognormal (-0.2342,0.35) |
| $1/\nu$ | lognormal(1.8318,0.6) |

Table S5: Prior distributions for *Manuka*

| Parameter | Prior distribution |
|---|---|
| $\beta_{11}$ | uniform(0.001,6) |
| $\beta_{12}$ | uniform(0.001,6) |
| $\beta_{22}$ | uniform(0.001,1.5) |
| $\beta_{21}$ | uniform(0.001,3) |
| $1/\delta(1-\omega)$ | lognormal (0.4039,0.6) |
| $1/\delta\omega$ | lognormal (0.4039,0.25) |
| $\tau$ | uniform(0.0001,1) |
| $1/\alpha(1-\tau)$ | lognormal (-0.2342,0.35) |
| $1/\nu$ | lognormal(1.8318,0.6) |
| $d$ | beta (1.5,30) |

##### S3.2 Summary statistics, distance criteria, and tolerance values of the ABC-SMC algorithm

***Medic:***

**Summary statistics:**

1. Daily new infections (confirmed influenza and unconfirmed influenza).
2. Total number of deaths throughout the epidemic by groups.
3. New infections from day 1 to 3.
4. Total number of infections throughout the epidemic by groups.
5. Number of healthy people removed on days 10 and 11.
6. Number infectious/ infected people removed on days 8, 9, 12 and 14.

**Distance criteria:**

**D1** Euclidean distance from the generated and observed new infections.

**D2, D3** For crew and passengers, the absolute difference between generated and observed total number of deaths. Absolute differences are calculated for both groups.

**D4, D5, D6** Absolute difference of generated and observed new infections on days 1, 2 and 3. Altogether, three distance criteria for the three days will be calculated.

**D7, D8** Absolute differences between generated and observed total number of infections throughout the epidemic for both crew and passengers.

**D9** Euclidean distance from the generated and observed number of removal of healthy people.

**D10** Euclidean distance from the generated and observed number of removal of infectious/ infected people.

**ABC-SMC pre-defined settings:**

The ABC-SMC algorithm was run through 5 generations to obtain 1000 samples from all the posterior distributions.

Table S6: Tolerance values for *Medic*

| | **Tolerance values through generations of ABC-AMC** | | | | |
|---|---|---|---|---|---|
| **Distance criteria** | **1** | **2** | **3** | **4** | **5** |
| D1 | 100 | 70 | 60 | 55 | 50 |
| D2 | 3 | 3 | 3 | 3 | 3 |
| D3 | 10 | 10 | 10 | 10 | 10 |
| D4, D5, D6 | 2 | 2 | 2 | 2 | 2 |
| D7 | 50 | 45 | 40 | 40 | 35 |
| D8 | 100 | 80 | 75 | 70 | 65 |
| D9 | 154 | 100 | 40 | 35 | 30 |
| D10 | 150 | 100 | 60 | 55 | 50 |

***Boonah:***

**Summary statistics:**

1. Total new infections throughout the epidemic by groups.
2. Daily new infections among the passengers.
3. Total number of deaths in both groups including deaths that occurred at the quarantine station as well as on board the ship.
4. Daily new cases in the first 5 days after leaving the source of infection.
5. Removal of infectious/ infected passengers on days from days 18 to 25 and day 29.
6. Sum of removal of healthy passengers from days 18 to 26.

7. Sum of removal of healthy passengers from days 21 to 31.
8. Sum of removal of the healthy, infectious, or infected crew from days 18 to 26.
9. Deaths that occurred in both groups on board the ship.

**Distance criteria:**

- **D1, D2** For crew and passengers, the absolute difference between generated and observed total new infections.
- **D3** Euclidean distance from the generated and observed infections among the passengers.
- **D4** Absolute difference between the observed and generated total number of deaths of both groups on board and at the quarantine station.
- **D5:D9** Absolute difference between the observed and generated number of infections from day 1 to 5 among the passengers.
- **D10** Euclidean distance from the observed and generated removal of infectious/ infected passengers on days from days 18 to 25 and day 29.
- **D11** Euclidean distance from the observed and generated removal of healthy people from summary statistics 6 and 7.
- **D12** Absolute difference between the observed and generated summary statistic 8.
- **D13** Absolute difference between the observed and generated total number of deaths of both groups on board.

**ABC-SMC pre-defined tolerance settings:**

The ABC-SMC algorithm was run on 3 generations to obtain 1000 samples were obtained from the posterior distributions.

Table S7: Tolerance values used in the ABC-SMC algorithm for *Boonah*

| | **Tolerance values through generations** | | |
|---|---|---|---|
| **Distance criteria** | **1** | **2** | **3** |
| **D1** | 35 | 25 | 20 |
| **D2** | 400 | 300 | 250 |
| **D3** | 115 | 100 | 80 |
| **D4** | 17 | 15 | 10 |
| **D5:9** | 6 | 6 | 6 |
| **D10** | 184 | 150 | 120 |
| **D11** | 149 | 120 | 100 |
| **D12** | 77 | 70 | 60 |
| **D13** | 10 | 8 | 6 |

***Devon:***

**Summary statistics:**

1. Daily new infections among the passengers.
2. Daily new infections among the crew.

**Distance criteria:**

- **D1** Euclidean distance from the generated and the observed infections among the crew.
- **D2** Euclidean distance from the generated and the observed infections among the passengers.

**ABC-SMC pre-defined tolerance settings:**

The ABC-SMC algorithm ran through 4 generations to obtain 1000 samples from posterior distributions.

Table S8: Tolerance values used in the ABC-SMC algorithm used for *Devon*

| | **Tolerance values through generations** | | | |
|---|---|---|---|---|
| **Distance criteria** | **1** | **2** | **3** | **4** |
| **D1** | 5 | 4 | 3 | 3 |
| **D2** | 31 | 28 | 26 | 24 |

***Manuka:***

**Summary statistics:**

1. Total number of infections among crew and passengers.
2. Total number of cases up to day 7.
3. Daily new infections that took place from day 8 onwards
4. Total number of deaths among both crew and passengers.

**Distance criteria:**

**D1, D2** For crew and passengers, the absolute difference between generated and observed total new infections.

**D3** Absolute difference between the observed and generated total number of cases up to day 7.

**D4** Euclidean distance from the generated and observed infections that took place from day 8 onwards (both crew and passengers together).

**D5** Absolute difference between the observed and generated total number of deaths.

**ABC-SMC pre-defined tolerance values**

Table S9: Tolerance values used in the ABC-SMC algorithm for *Manuka*

| | **Tolerance values through generations** | | | |
|---|---|---|---|---|
| **Distance criteria** | **1** | **2** | **3** | **4** |
| **D1** | 25 | 18 | 10 | 8 |
| **D2** | 8 | 7 | 6 | 5 |
| **D3** | 25 | 18 | 15 | 12 |
| **D4** | 7 | 6 | 5 | 4 |
| **D5** | 3 | 2 | 1 | 1 |

#### S4 Hierarchical analysis

##### S4.1 Conditional re-sampled paths and re-sampled paths

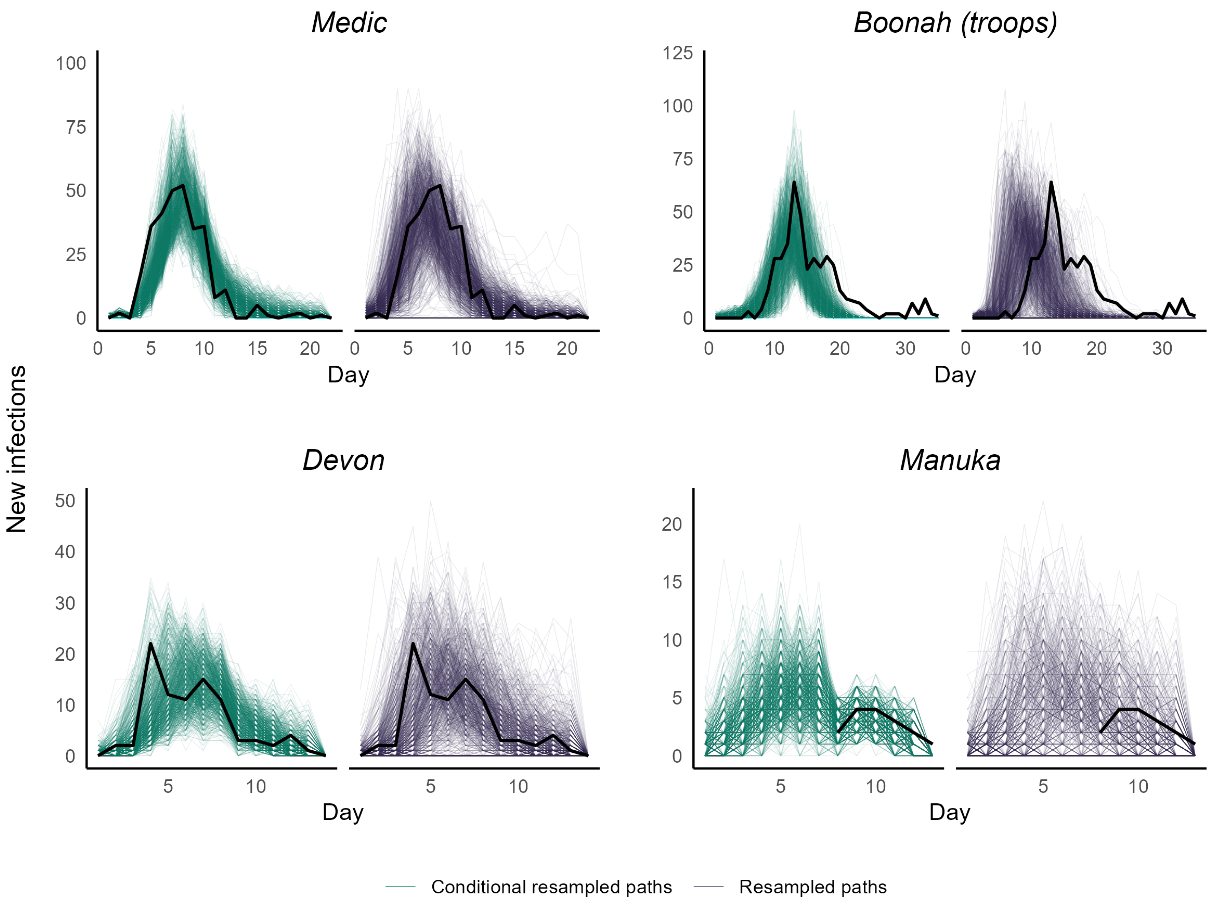


Figure S10: Conditional re-sampled paths (in green) by the hierarchical estimation algorithm and re-sampled paths (in purple) by the estimated parameters. Black lines are the observed data.

##### S4.2 Hyper-parameters

We estimated the hyper-parameters $\Psi_{CC}, \Psi_{CP}, \Psi_{PC}, \Psi_{PP}, \sigma_{CC}, \sigma_{CP}, \sigma_{PC}, \sigma_{PP}$ within a pseudo-marginal setting. The approximate likelihood (see Alahakoon et al. (2022) for the details) at the hyper-parametric level, $\hat{p}(\mathbf{y}|\Psi)$ was

$$\hat{p}(\mathbf{y}|\Psi) \propto \prod_{k=1}^{4} \sum_{j=1}^{1000} \frac{p(\beta^{(j)}|\Psi)}{p(\beta^{(j)})},$$

where $\mathbf{y}$ is the observed data. $p(\beta^{(j)}|\Psi)$ is the conditional prior distribution and $p(\beta^{(j)})$ is the prior distribution for transmission rates. We used this likelihood within an MCMC framework and obtained 40000 samples from each of the marginal posterior distributions of the hyper-parameters. Then we used every 40th sample and obtained 1000 samples for the posterior distributions of the hyper-parameters.

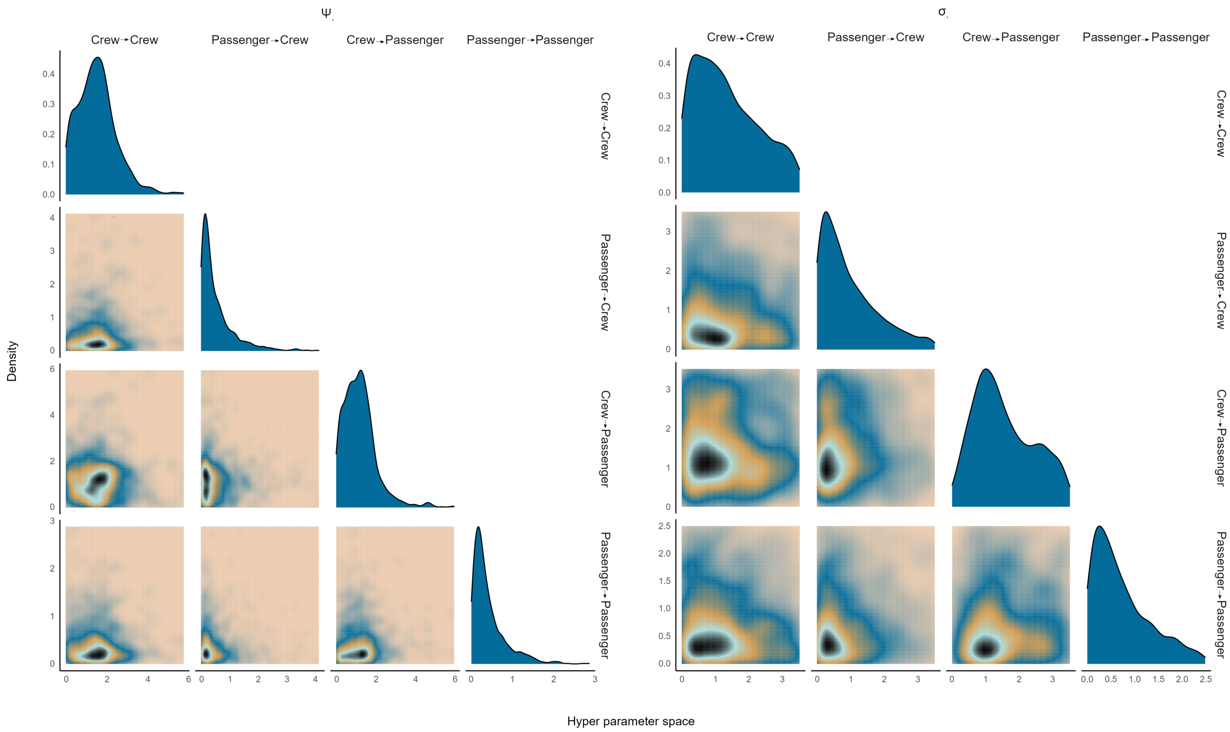


Figure S11: Posteriors for hyperparameters

##### S4.2.1 Comparison of transmission rates under a hierarchical analysis and an independent analysis

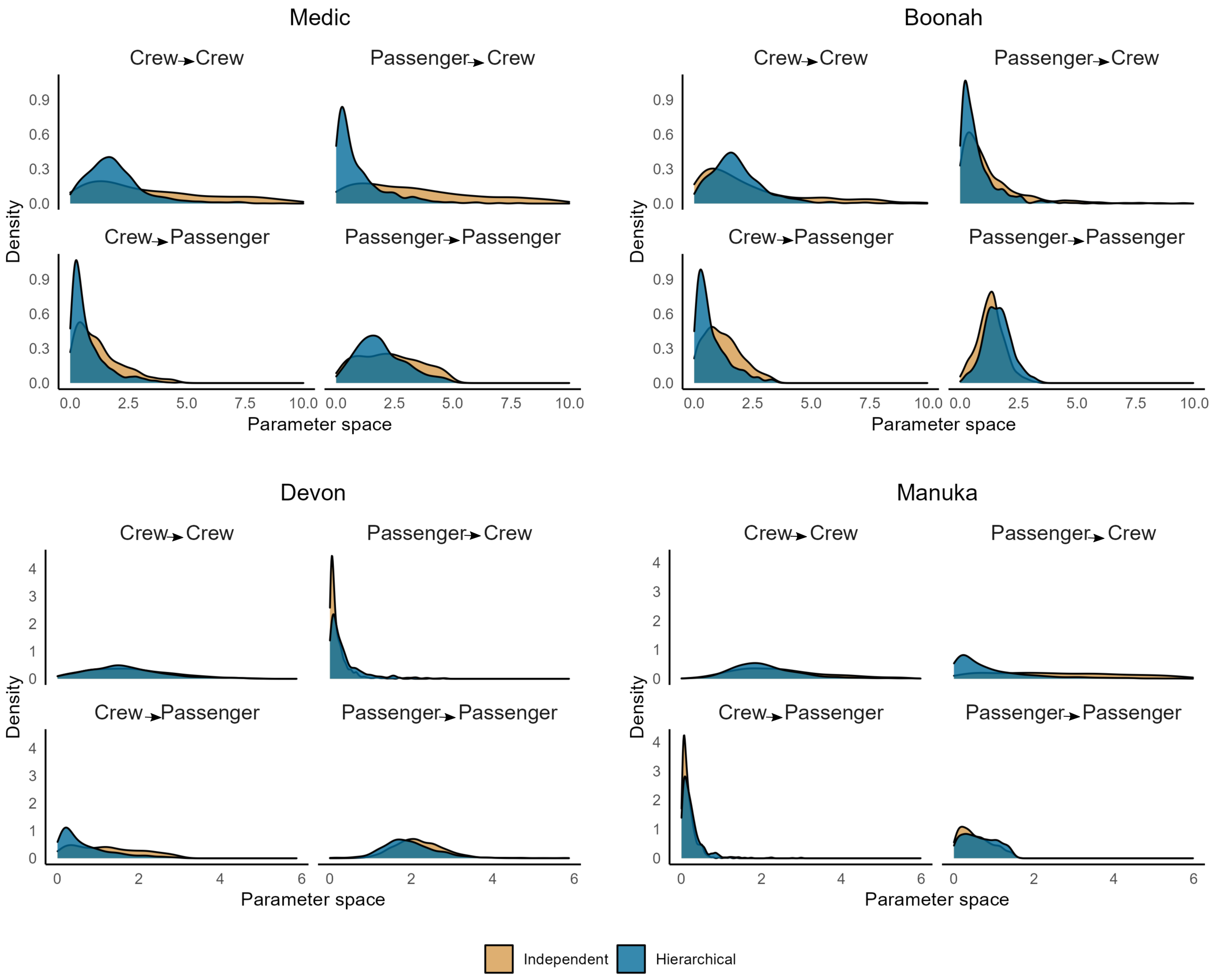


Figure S12: Comparison of transmission rates under hierarchical analysis and independent analysis

##### S4.3 Other parameter estimates under a hierarchical analysis

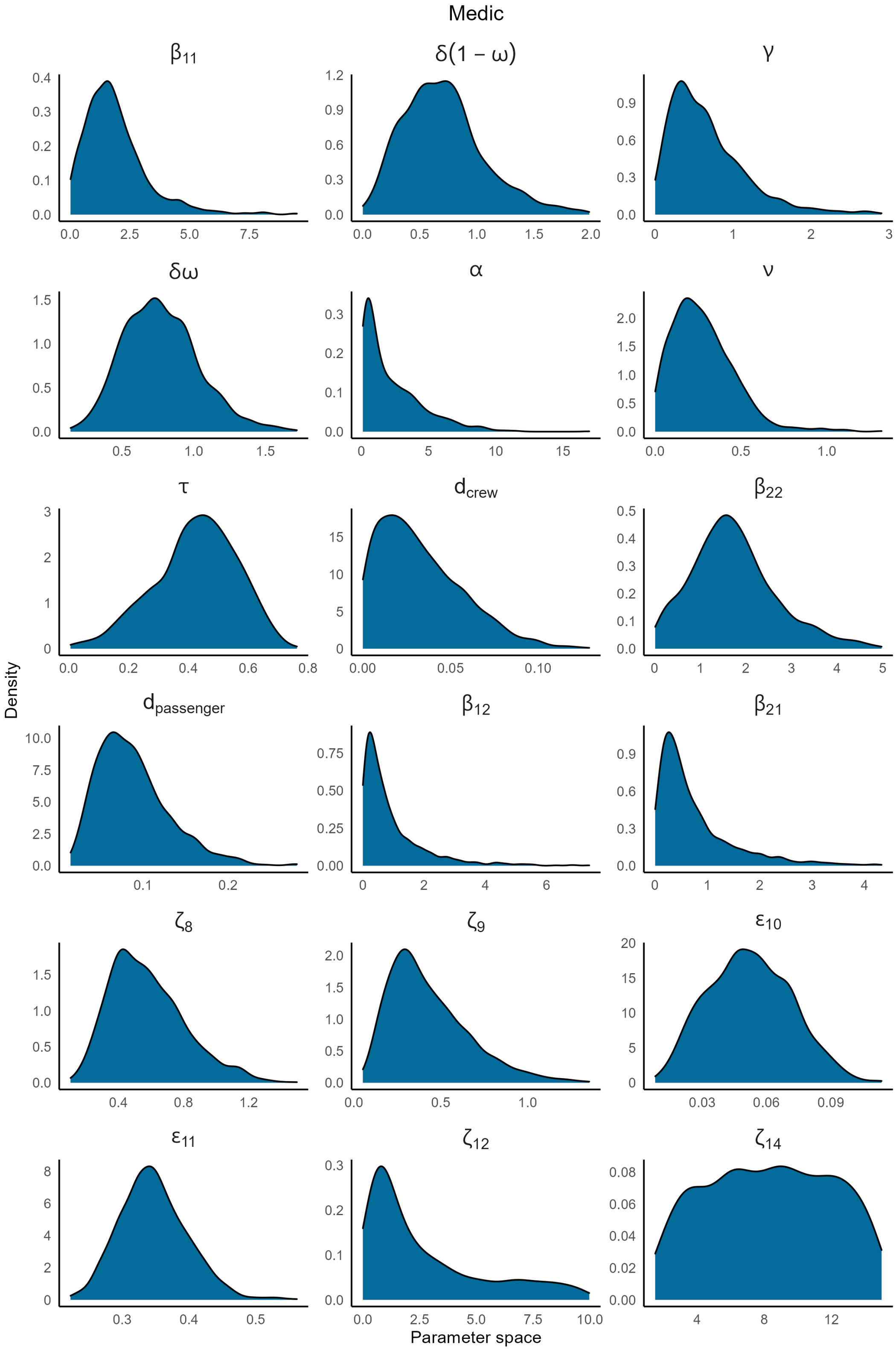


Figure S13: Posterior distributions for *Medic*

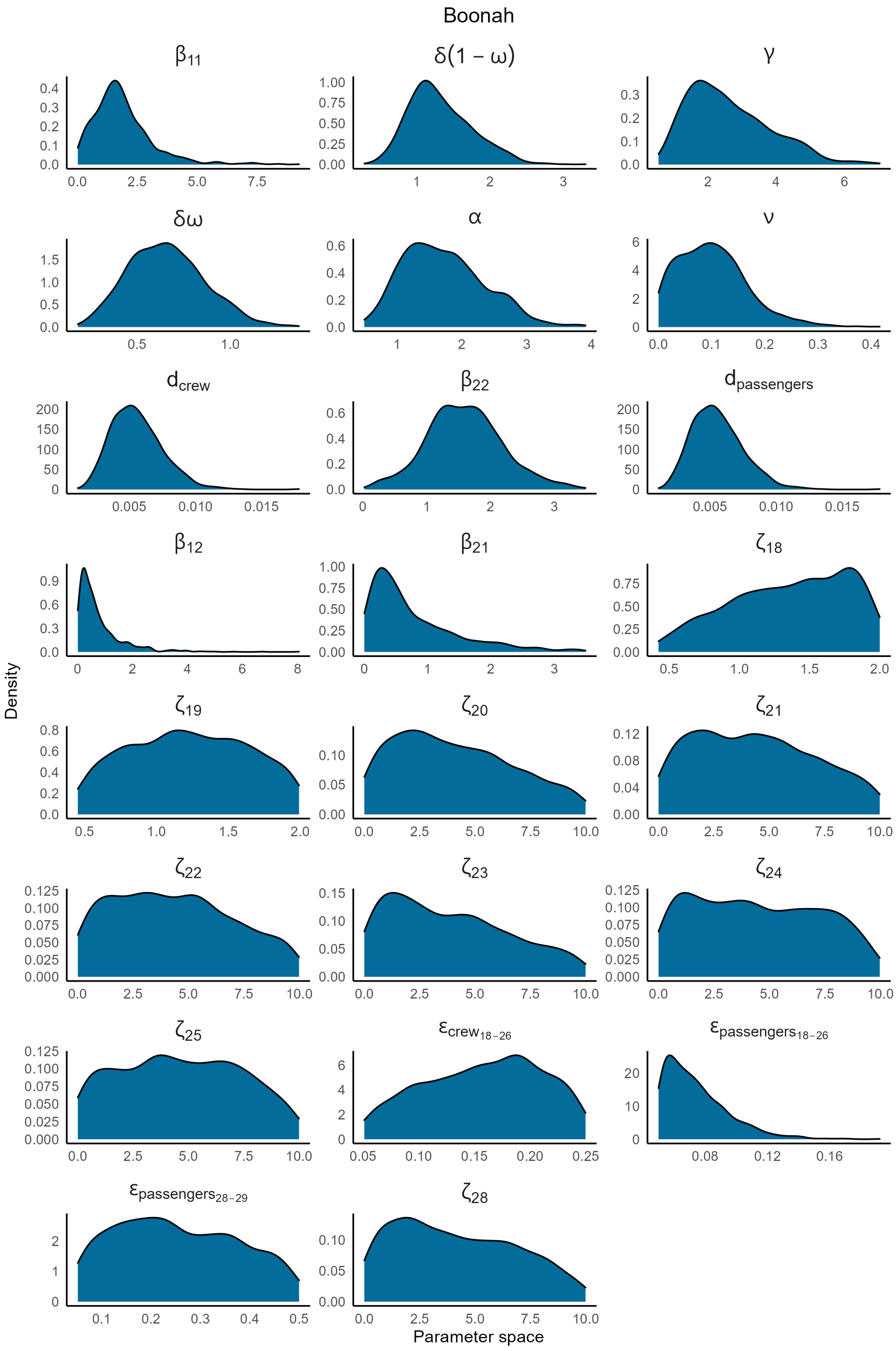


Figure S14: Posterior distributions for *Boonah*

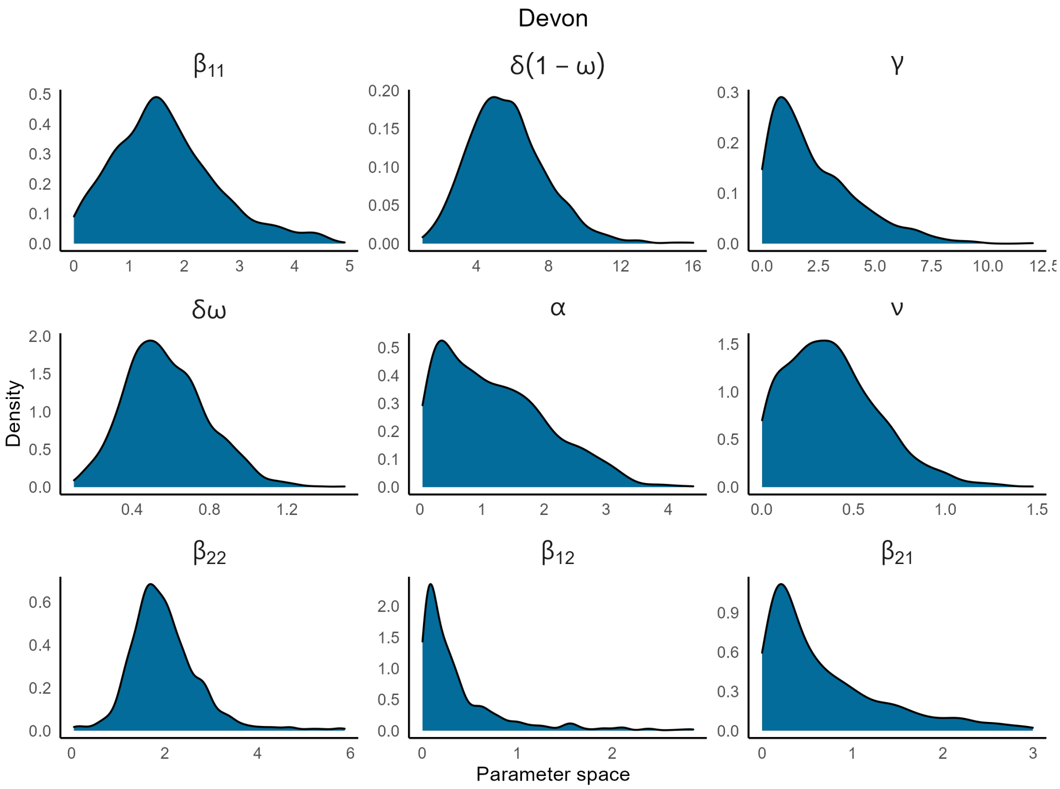


Figure S15: Posterior distributions for *Devon*

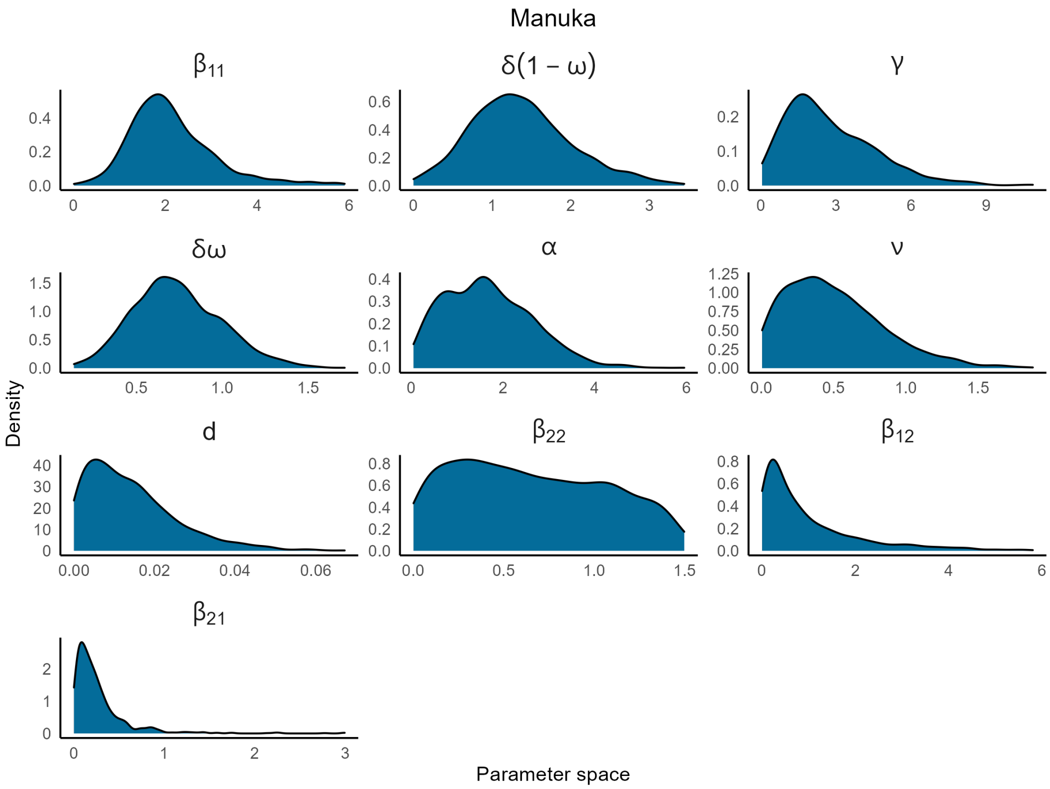


Figure S16: Posterior distributions for *Manuka*

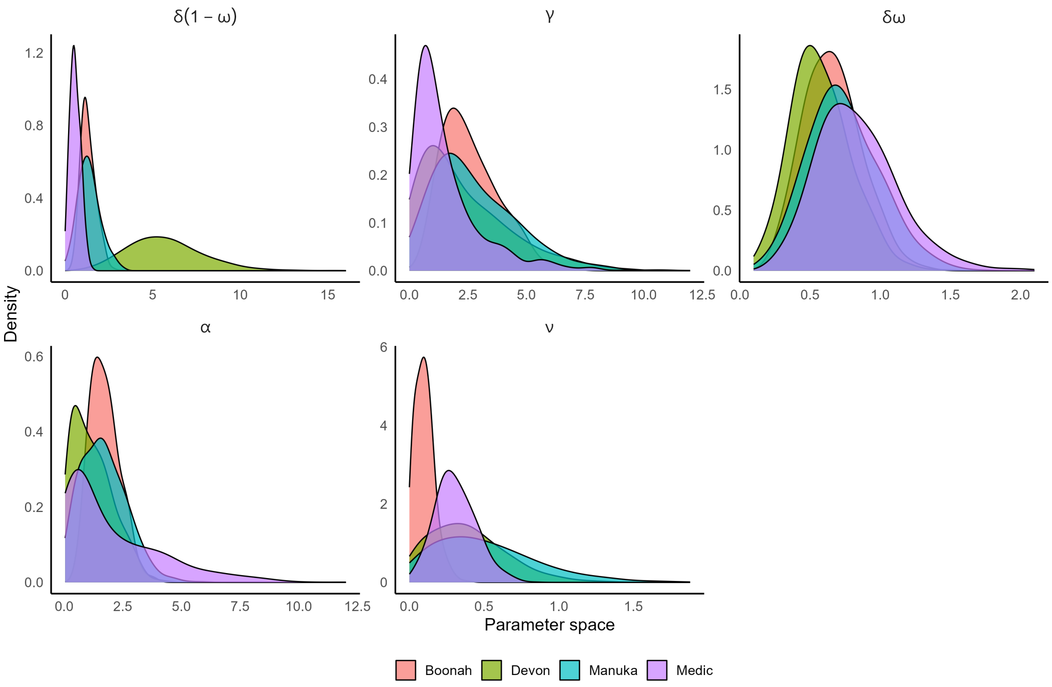


Figure S17: Overlay of posterior distributions of parameters that are common to all the outbreaks

#### S5 Removal of infectious/infected and healthy individuals based on re-sampled paths

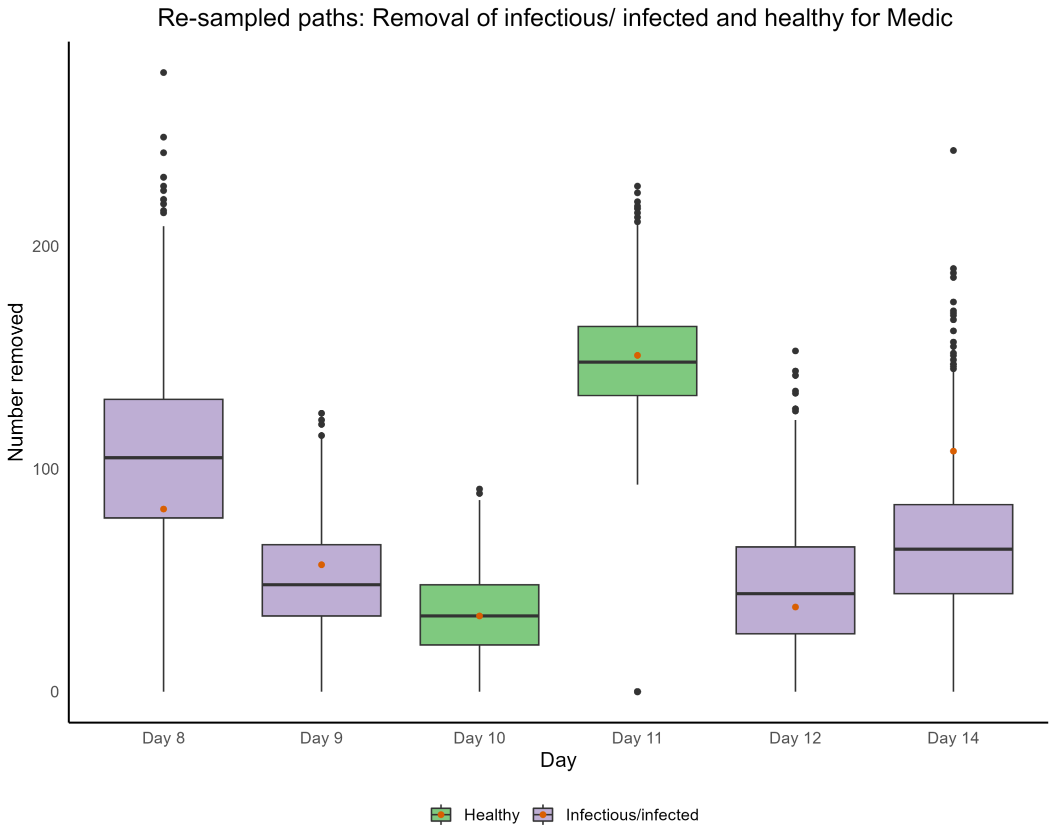


Figure S18: Removals estimated based on re-sampled paths for *Medic.* Box plots in purple are for infectious/infected individuals, and box plots in green are for healthy individuals. The orange dots are observed values from the data.

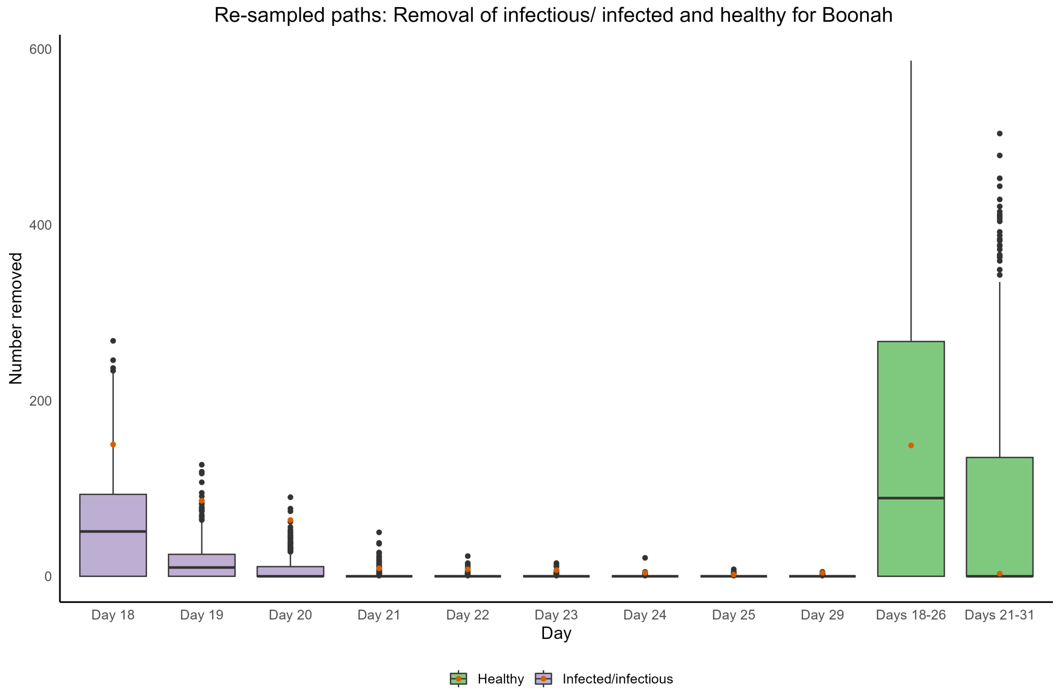


Figure S19: Removals estimated based on re-sampled paths for troops of *Boonah.* Box plots in purple are for infectious/infected individuals, and box plots in green are for healthy individuals. The orange dots are observed values from the data.

#### S6 Comparison of hierarchical and independent estimation based on re-sampled paths

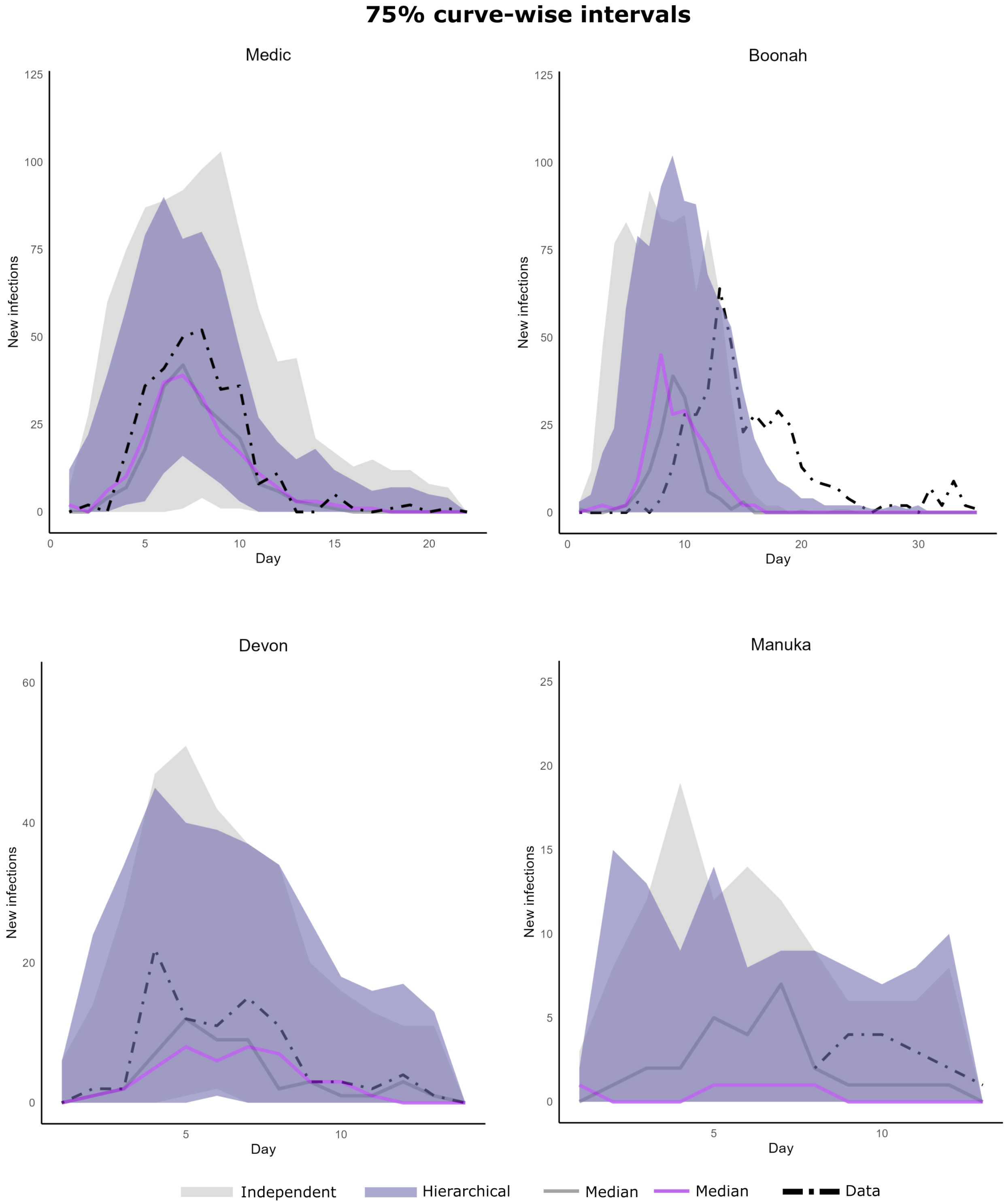


Figure S20: Comparison of independent and hierarchical estimation frameworks based on 75% curve-wise intervals.
Intervals in grey are for independent estimation and those in purple are for hierarchical estimation. Lines in grey and purple are median paths for independent and hierarchical estimation frameworks respectively. Black dashed lines are observed data.

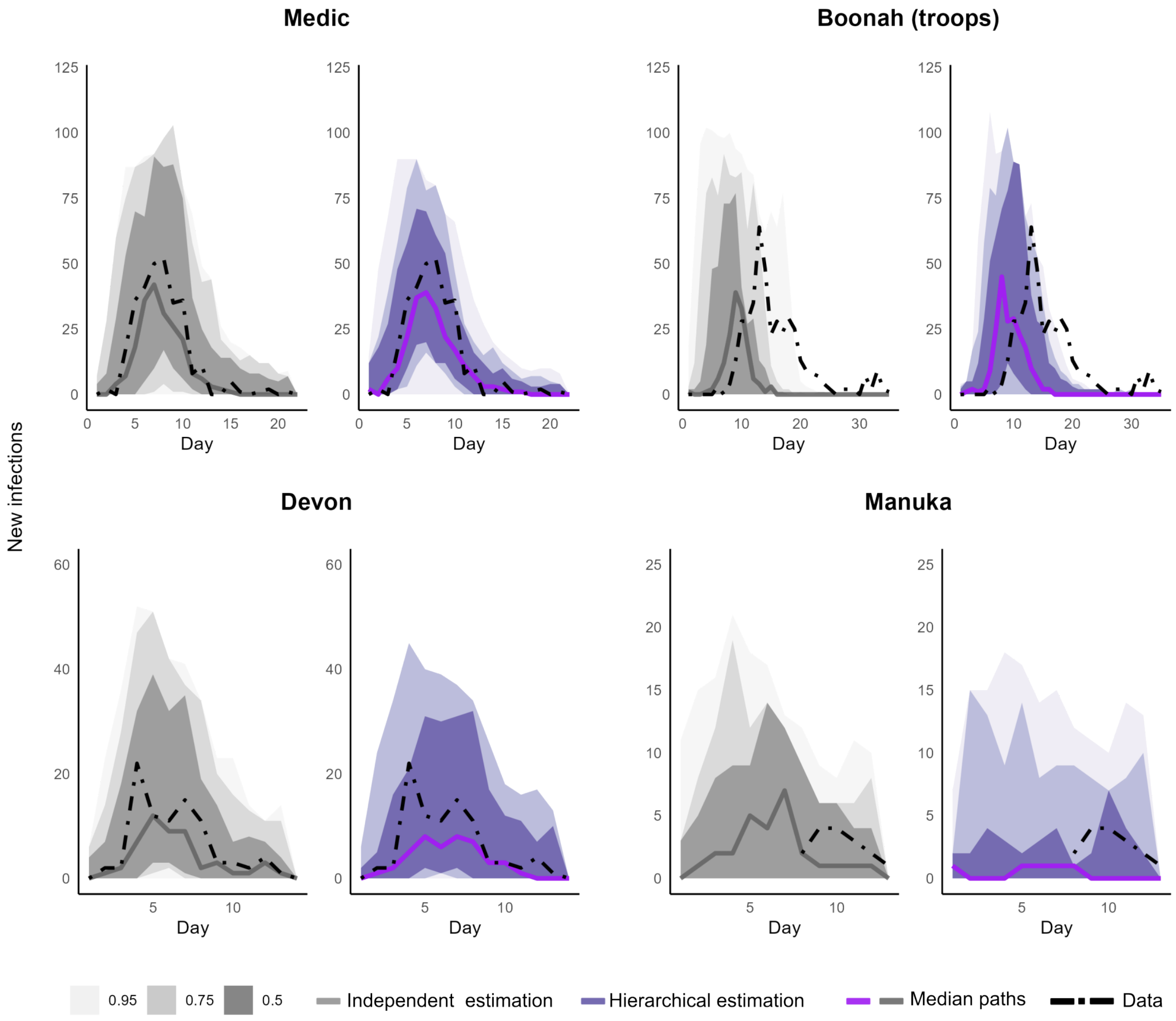


Figure S21: Comparison of independent and hierarchical estimation frameworks based on (50, 75, 95)% curve-wise intervals.
Intervals in grey are for independent estimation and those in purple are for hierarchical estimation. Lines in grey and purple are median paths for independent and hierarchical estimation frameworks respectively. Black dashed lines are observed data.

#### S7 Use of other model structures for *Medic*

To test whether other simple model structures were suitable for Medic we also undertook our analysis with a simple two-group SEAIR-type model in which we excluded the compartments for those classified as mild infectious ($M_i$), mild recovering ($C_{Mi}$), and severely infected and recovering ($C_i$)). This model failed to accurately predict the removal of infectious, infected, and healthy individuals (Figure S22).

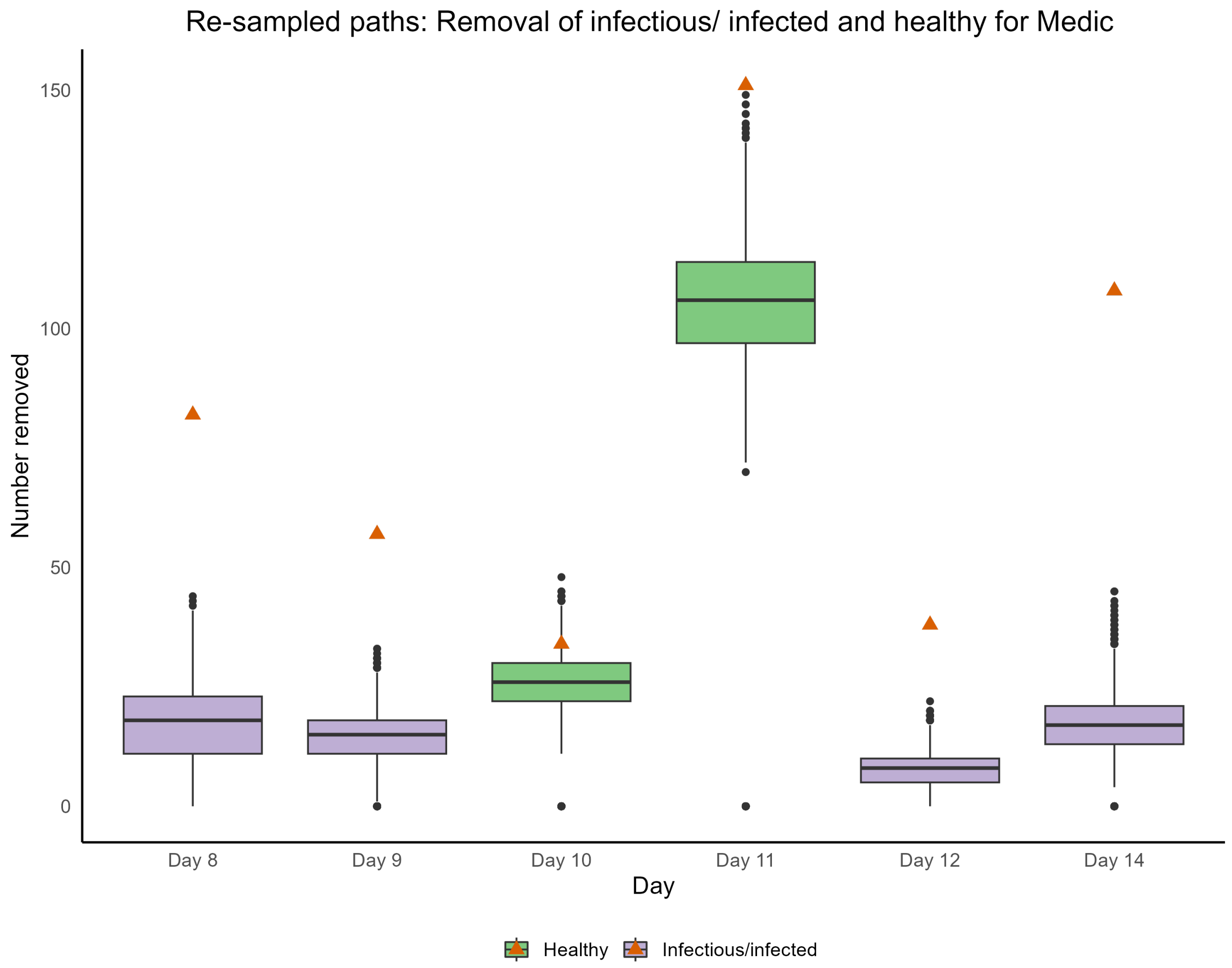


Figure S22: Boxplots of removal of infectious, infected and healthy individuals of *Medic* based on a simpler model that does not take into account compartments $M_i, C_{Mi}$, and $C_i$ based on re-sampled paths. The observed values are represented as orange triangles.

##### S7.1 MATLAB Codes

The codes can be found on GitHub at `https://github.com/PunyaAlahakoon/Ship_outbreaks_1918.git`

#### References

Punya Alahakoon, James M McCaw, and Peter G Taylor. Estimation of the probability of epidemic fade-out from multiple outbreak data. *Epidemics*, page 100539, 2022.

Tina Toni, David Welch, Natalja Strelkowa, Andreas Ipsen, and Michael PH Stumpf. Approximate bayesian computation scheme for parameter inference and model selection in dynamical systems. *Journal of the Royal Society Interface*, 6(31):187–202, 2009.
